# Biological age uncertainty reveals discordant aging and predicts human healthspan

**DOI:** 10.64898/2026.09.26.26364091

**Authors:** Yanjun Li, Guoqing Feng, Shouyi Yan, Qi Huang, Jin Jiang, Yufei Wang, Zeyuan Pei, Lyn Xuan Tay, Keliang Li, Zean Pan, Zhichao Yuan, Liangcai Gao, John S. Ji, Limei Ke, Kuiying Gu, Qian Di

## Abstract

Biological aging clocks typically summarize complex biological profiles as a single age estimate, yet how uncertain these estimates are and whether this uncertainty represents a distinct, biologically meaningful dimension of human aging remain unclear. Here, we develop a probabilistic framework across nuclear magnetic resonance (NMR) metabolomic, plasma proteomic and routine blood test (RBT) profiles in more than 450,000 UK Biobank participants to derive biological age gap (BAG) and biological age uncertainty (BAU) from the mean and width of predicted-age distributions. BAU was weakly correlated with BAG and increased with age-signal disagreement across individual molecules and organs, linking uncertainty to discordant aging across biological systems. Higher BAU predicted mortality, loss of healthspan and multimorbidity independently of BAG, with mortality hazard ratios of 1.24 (95% CI, 1.23–1.26) and 1.40 (95% CI, 1.37–1.43) per standard deviation for NMR- and proteomic-derived BAU, respectively. Adding BAU to conventional clinical models improved 10-year risk discrimination; moreover, longitudinal increases in NMR-derived BAU tracked subsequent health decline, and mortality associations were replicated using RBT-derived BAU across three independent external cohorts. Multi-omics and physiological analyses linked BAU to inflammation, metabolic dysregulation and tissue remodeling beyond BAG. Genetic analyses further revealed overlapping but distinct architectures of BAU and BAG, with BAU showing stronger genetic correlations with frailty and cardiometabolic diseases, and tissue and functional mapping implicating hepatic, immune and lipid-metabolic processes. This study establishes a framework for characterizing biological age through both its mean deviation and uncertainty, defining BAU as an independent dimension of human aging that links discordant aging to future healthspan.

## Introduction

Aging is a heterogeneous process. Individuals of the same chronological age differ substantially in physiological decline, molecular trajectories, and risks of aging-related disease and death^1–4^. Biological aging clocks made these differences quantifiable by estimating age from clinical, epigenetic and other molecular profiles^5–7^. Organ- and system-specific clocks subsequently revealed distinct aging trajectories among organs and physiological systems within the same individual^1,2,8–10^. Recently, tissue-^11^, cell-type-^12^, and transcriptomic clocks^13^ have extended this resolution further. Therefore, aging can now be resolved across a nested hierarchy, from variation between individuals to variation among the organs, tissues, cell types, and molecular programs within them. However, across this hierarchy, aging clocks typically summarize a multidimensional biological profile as a single estimated age^3,14^.

The difference between this single estimate and chronological age, termed the biological age gap (BAG), quantifies age deviation and is associated with healthspan, aging-related disease and mortality^3,7,15–17^. Nonetheless, biological age estimates carry predictive uncertainty, since a single biological profile often corresponds to a distribution of potential biological aging. The breadth of this distribution indicates the precision of the estimate^14,18^. To make this uncertainty explicit, recent studies have applied individualized prediction intervals to epigenetic clocks during childhood development^19^. Building on this observation, we define the width of an individual predictive age distribution as biological age uncertainty (BAU) and hypothesize that BAU represents a new dimension of biological aging, independent of BAG.

Biologically, this uncertainty reflects how consistently the signals within an individual profile support a common aging state. Across physiological systems, aging progressively weakens functional coupling^20–22^, eroding the coordinated regulation required to preserve physiological integrity^23^. Intuitively, the divergence among aging signals captured by BAU can therefore serve as a proxy for discordant aging across biological systems. Thus, it remains unclear whether BAU captures an independent dimension of human biological aging distinct from the age deviation quantified by BAG, and whether this uncertainty reflects discordant aging across biological systems and marks vulnerability to future health decline.

Here, we developed a probabilistic modelling framework that represents biological age as an individual predictive distribution, jointly characterizing age deviation and uncertainty. Within this framework, BAG and BAU are derived from the mean and width of the same distribution, respectively. We applied this framework to large-scale nuclear magnetic resonance (NMR) metabolomic and plasma proteomic profiles from UK Biobank. We first evaluated whether BAU represents a dimension of biological aging distinct from BAG and whether higher BAU is associated with greater disagreement among molecular age signals and organ-specific BAGs. We then examined whether baseline BAU and changes in BAU between assessments were associated with subsequent mortality, loss of healthspan, multimorbidity and incident diseases after accounting for BAG. We also assessed whether adding BAU to conventional clinical risk models improved predictive discrimination. To evaluate generalizability, we developed models based on routine blood tests (RBT) and evaluated the associations of RBT-derived BAU with mortality in three independent external cohorts. We compared the molecular and pathway associations of BAU with those of BAG and examined whether these differences were reflected in physiological and clinical traits. Finally, we characterized the genetic architectures of BAG and BAU, examined their genetic correlations with aging-related traits, and used tissue and functional mapping to identify associated tissues, cell types and biological pathways. Taken together, these analyses demonstrate that biological aging can be characterized by both mean age deviation and uncertainty in the age estimate, with greater uncertainty potentially reflecting more discordant aging across biological systems.

## Results

### BAU is distinct from BAG

We developed a probabilistic modelling framework to derive BAG and BAU, with a mathematical formulation providing a theoretical interpretation of these measures in terms of mean aging and discordant aging across biological subsystems (Fig. 1a, Methods, and Supplementary Notes 1-4). Separate NMR and proteomic age models were trained and independently calibrated in healthy reference participants, then applied to participants in held-out assessment centres. This yielded predicted-age distributions for 272,672 participants with NMR measurements and 43,867 with proteomic measurements (Extended Data Fig. 1). BAG represented the deviation of mean predicted age from chronological age, whereas BAU represented the width of the predicted-age distribution (Fig. 1b). Both measures were derived from calibrated predictions, with BAU calculated from log predictive variance, and were adjusted for age and standardized within sex relative to healthy reference participants.

**Fig. 1.**
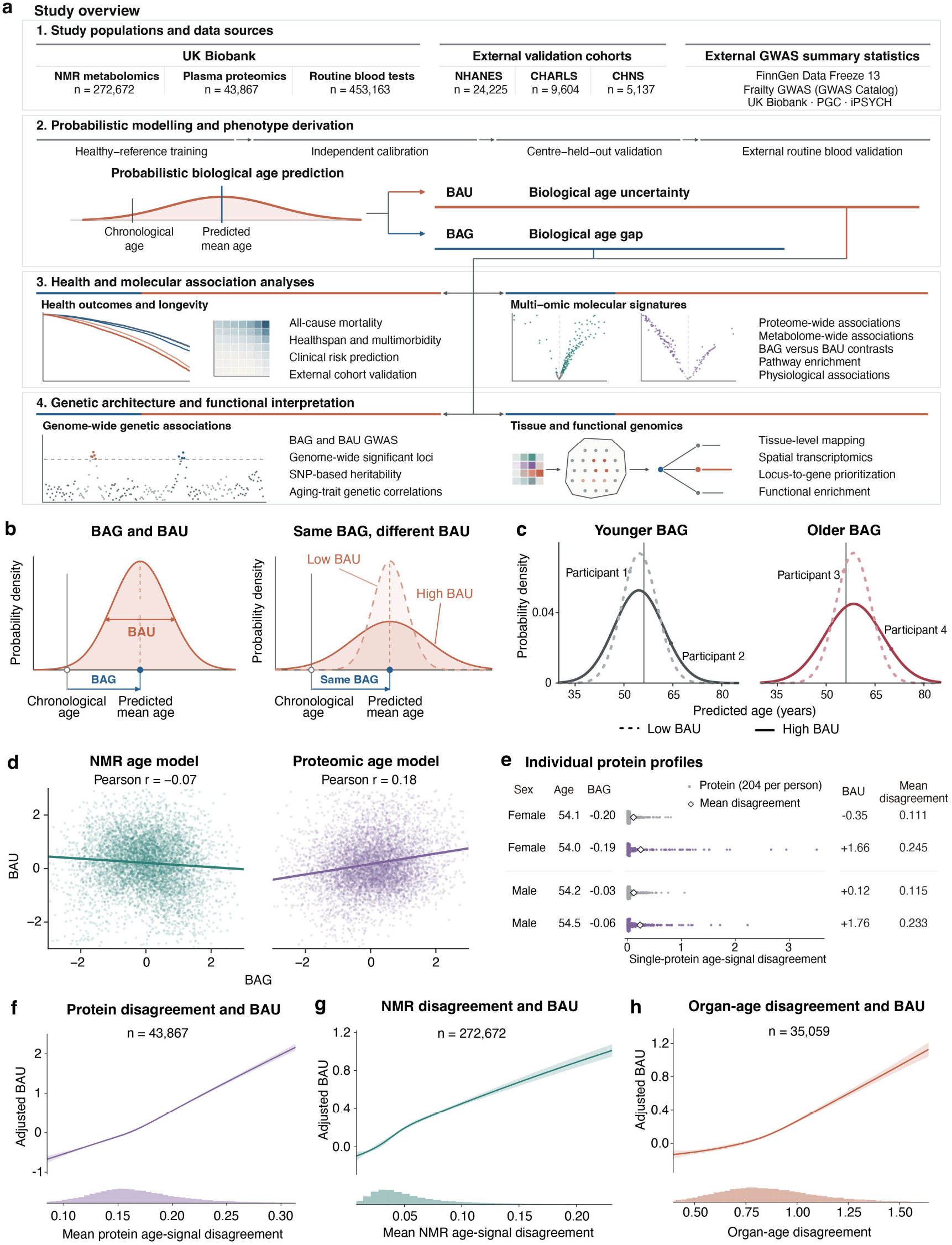
BAU captures a distinct dimension of biological aging. **a,** Study overview. UK Biobank NMR metabolomic profiles, plasma proteomic profiles and routine blood tests (RBT) were used to develop probabilistic age models through healthy-reference training, independent calibration and validation in held-out assessment centres. RBT models were further validated in three external cohorts. BAG and BAU were examined in relation to health outcomes, molecular profiles and genetic variation, followed by tissue mapping and functional analyses. **b,** Conceptual illustration of BAG and BAU. BAG describes the deviation of mean predicted age from chronological age, whereas BAU describes the width of the predicted age distribution. Individuals with the same BAG can have different BAU, illustrated by narrower and broader distributions. **c,** NMR-derived predicted-age distributions for two pairs of participants matched for age, sex and BAG, with different BAU. Dashed and solid curves indicate lower and higher BAU, respectively. Grey vertical lines mark chronological age. **d,** Relationships between BAG and BAU derived from NMR and proteomic models. Lines show linear regression fits, with Pearson correlation coefficients indicated. **e,** Protein age-signal disagreement in two pairs of participants matched for age, sex and BAG. Each row represents one participant. For each protein, likelihood measures how well the participant’s protein level matches the levels expected at a given age in the healthy reference population. The age with the highest mean log-likelihood across all 204 proteins was selected as the common support age. Each dot shows the decrease from a protein’s maximum log-likelihood to its log-likelihood at this common age. Larger values indicate greater disagreement between that protein’s age signal and the common support age. Diamonds show mean disagreement across proteins (Extended Data Fig. 2a). **f,** Association between mean age-signal disagreement across 204 proteins and proteomic-derived BAU (n = 43,867). The curve shows mean BAU adjusted for BAG, age, sex, assessment centre and measurement missingness; shading indicates the 95% confidence interval. The histogram shows the distribution of disagreement. **g,** As in **(f)**, for age-signal disagreement across 168 NMR measures and NMR-derived BAU (n = 272,672). **h,** As in **(f)**, for organ-age disagreement and proteomic-derived BAU (n = 35,059), with additional adjustment for organ-panel missingness. Organ-age disagreement is the within-participant standard deviation of standardized BAG across ten organ clocks. NMR, nuclear magnetic resonance; RBT, routine blood tests.

We first examined whether BAU distinguished individuals with similar BAG. Participants matched for age, sex and NMR-derived BAG had predicted-age distributions with similar means but different widths (Fig. 1c). Across the study populations, BAG and BAU were weakly correlated for both NMR-derived measures (Pearson r = −0.07) and proteomics-derived measures (r = 0.18; Fig. 1d). These observations indicate that BAG and BAU captured distinct aspects of the predicted age distribution.

Individual molecular measurements within the same person can support different ages when compared with healthy reference profiles. We therefore examined whether greater disagreement among these molecular age signals was associated with higher BAU. Using healthy reference profiles, we identified the age that best fitted each participant’ s molecular measurements collectively. For each molecule, we calculated the difference in log-likelihood between its individually best-fitting age and this common age, then averaged these differences to quantify molecular age-signal disagreement (Extended Data Fig. 2a). Among participants with similar BAG, greater protein age-signal disagreement accompanied higher proteomic-derived BAU (Fig. 1e). Across both assays, greater molecular age-signal disagreement was associated with higher BAU after adjustment for BAG and demographic and assay covariates (Fig. 1f,g).

We next tested whether this relationship extended to differences in age estimates across organs. We quantified organ-age disagreement as the within-participant SD of ten proteomic-derived organ BAGs (Extended Data Fig. 2b,c). Greater organ-age disagreement was associated with higher proteomic-derived BAU after adjustment for BAG and demographic and assay covariates (Fig. 1h). Thus, the association between BAU and disagreement among age signals was evident both among individual molecular measurements and among organ age estimates.

We further evaluated whether BAU was consistent over time, whether measured potential confounders explained its variation, and whether its estimates depended on algorithm choice. Baseline and repeat NMR-derived BAU were moderately correlated (Spearman r = 0.47, as for BAG), indicating that higher BAU tended to persist across assessments (Extended Data Fig. 2d). Models incorporating age, sex, BAG and technical, lifestyle, socioeconomic, medication and baseline clinical factors explained 27.2% of NMR-derived BAU variation and 23.7% of proteomic-derived BAU variation. After subtracting these models’ predictions, the remaining BAU was strongly correlated with original BAU (Spearman r = 0.843 and 0.854, respectively), indicating that the measured factors accounted for a limited proportion of BAU variation and that higher BAU remained evident after adjustment (Extended Data Fig. 2e). BAU estimates were also strongly correlated across three probabilistic algorithms, indicating that participants with higher BAU were consistently identified across model choices (Extended Data Fig. 2f). Together, these findings support the consistency of BAU over time and across algorithms, with its variation not primarily explained by measured potential confounders.

Collectively, these results characterize BAU as a robust phenotype distinct from the mean age deviation measured by BAG. Higher BAU reflects a broader predicted-age distribution and is associated with greater disagreement among molecular and organ age signals.

### BAU stratifies future health risk beyond BAG

We next examined whether BAU stratified future health risk beyond BAG. Participants were classified into four groups defined by low or high BAG and BAU, using zero on the healthy-reference scale as the threshold. Over 15 years, participants with high BAU had higher cumulative probabilities of all-cause mortality, loss of healthspan and major disease multimorbidity within both low and high BAG groups in both proteomic- and NMR-based models (Fig. 2a,c). In models including BAG and BAU together and adjusted for demographic, behavioural and socioeconomic covariates, each 1 SD increase in NMR-derived BAU was associated with increased mortality (HR, 1.242; 95% CI, 1.226 to 1.258), loss of healthspan (HR, 1.151; 1.139 to 1.162) and multimorbidity (HR, 1.212; 1.193 to 1.231). The corresponding hazard ratios for proteomics-derived BAU were 1.399 (1.368 to 1.431), 1.238 (1.207 to 1.270) and 1.289 (1.244 to 1.336), respectively (Fig. 2b,d).

**Fig. 2.**
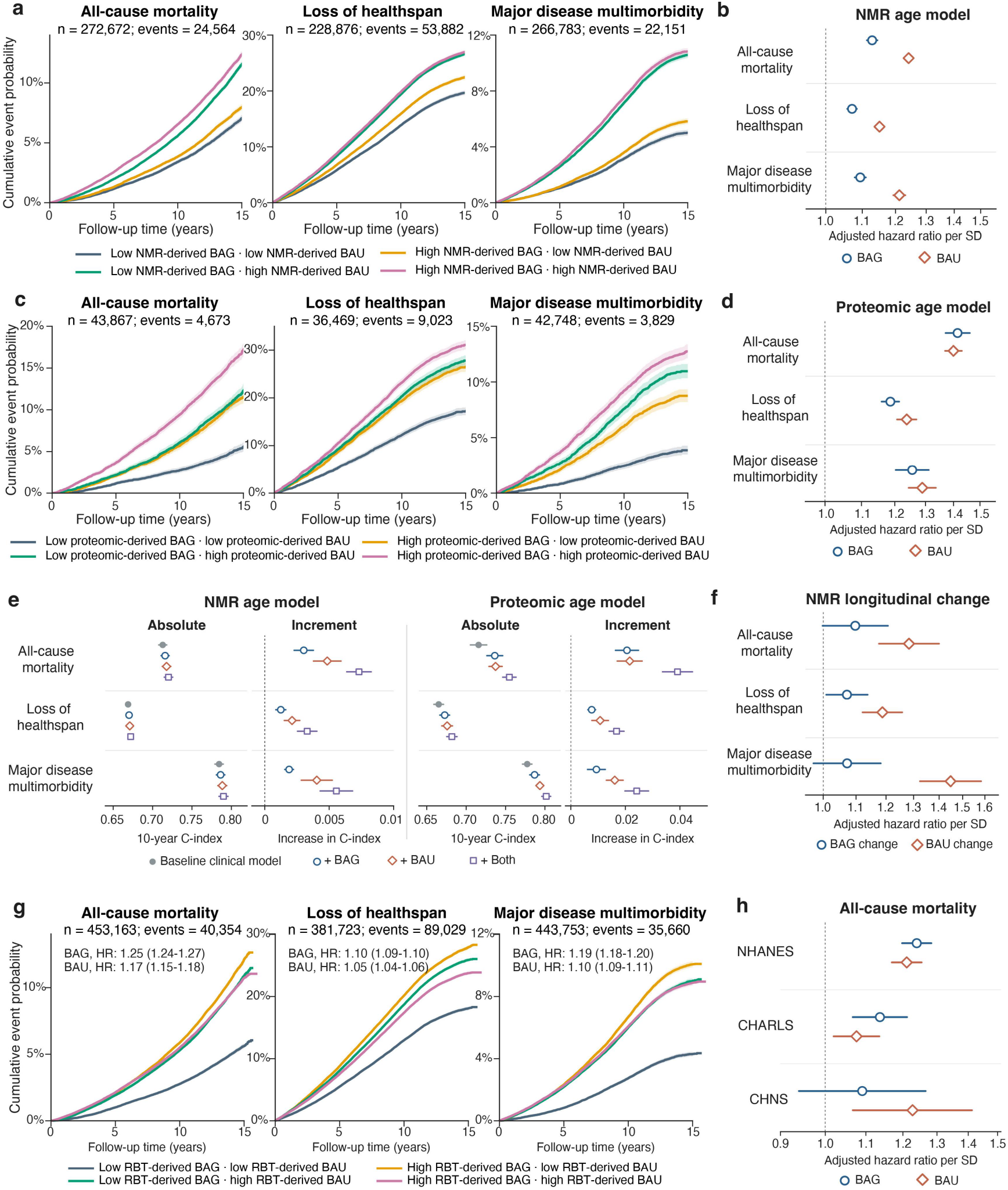
BAU stratifies future health risk beyond BAG. **a,** Fifteen-year cumulative probabilities of all-cause mortality, loss of healthspan and major disease multimorbidity in four groups defined by low or high nuclear magnetic resonance (NMR)-derived BAG and BAU. Low and high values were defined as <0 and ≥0 on the healthy-reference scale, respectively. Loss of healthspan comprised the first major disease or death, and multimorbidity the occurrence of a second distinct major disease domain. Curves show one minus Kaplan-Meier estimates for mortality and loss of healthspan, and Aalen-Johansen cumulative incidence for multimorbidity, accounting for competing mortality. Shading indicates 95% confidence intervals. **b,** Associations of NMR-derived BAG and BAU with the three outcomes. Hazard ratios (HRs) are reported per 1 SD from Cox models including BAG and BAU together and adjusted for demographic, behavioural and socioeconomic factors. Points and horizontal lines show HRs and 95% confidence intervals. **c,d,** As in **(a,b),** for proteomic-derived BAG and BAU. **e,** Ten-year C-index and its increase after adding BAG, BAU or both to a baseline clinical model, shown separately for NMR-derived and proteomic-derived measures. The baseline model included age, sex, smoking, diabetes history, systolic blood pressure, total cholesterol, HDL cholesterol and antihypertensive treatment. Increase in C-index is the difference between each expanded model and the baseline model. Predictions were evaluated in five folds with separate assessment centres used for training and validation. Points and horizontal lines show estimates and 95% confidence intervals. **f,** Associations of changes in NMR-derived BAG and BAU with subsequent health outcomes. Change was calculated as the value at reassessment minus the baseline value, and follow-up began at reassessment. Cox models included baseline BAG, BAU, both change measures, the interval between assessments, demographic, behavioural and socioeconomic factors. Points and horizontal lines show HRs per 1 SD of change and 95% confidence intervals. **g,** As in **(a)**, for RBT-derived BAG and BAU in UK Biobank. Values within each plot are adjusted HRs and 95% confidence intervals per 1 SD from Cox models including BAG and BAU together. **h,** Associations of RBT-derived BAG and BAU with all-cause mortality in NHANES (n = 24,225; 3,003 deaths), CHARLS (n = 9,604; 806 deaths) and CHNS (n = 5,137; 142 deaths). UK Biobank models using the blood measurements available in each cohort were applied with the UK Biobank calibration and healthy-reference scale. Cox models included BAG and BAU together and cohort-specific demographic and health covariates. Points and horizontal lines show HRs per 1 SD and 95% confidence intervals. NMR, nuclear magnetic resonance; RBT, routine blood tests.

We then assessed whether BAU improved risk discrimination when added to conventional clinical factors. In held-out assessment centres, adding BAU to the baseline clinical model increased the 10-year C-index by 0.002 to 0.005 for NMR and 0.011 to 0.022 for proteomics across the three primary outcomes, respectively. Models combining both BAG and BAU achieved the largest improvement for each outcome, with increments of 0.003 to 0.007 for NMR and 0.017 to 0.039 for proteomics (Fig. 2e). BAU was also associated with nine incident diseases spanning cardiovascular, metabolic, renal, respiratory, neurological and cancer outcomes, with hazard ratios ranging from 1.11 to 1.47 for NMR and 1.18 to 1.49 for proteomics (Extended Data Fig. 3a,b). Models combining BAG and BAU improved discrimination over the baseline clinical model for all nine diseases in both proteomic- and NMR-based models (Extended Data Fig. 3c,d).

Longitudinal changes in BAU also carried information about subsequent health. Among 13,620 participants with repeat NMR measurements, we assessed whether BAU change between reassessment and baseline was associated with health outcomes. Each 1 SD higher BAU change was associated with subsequent mortality (HR, 1.283; 95% CI, 1.176 to 1.399), loss of healthspan (HR, 1.187; 1.121 to 1.258) and multimorbidity (HR, 1.448; 1.324 to 1.583), after accounting for baseline BAG and BAU, BAG change and demographic, behavioural and socioeconomic covariates (Fig. 2f). Associations between BAU change and the nine incident diseases were also positive (Extended Data Fig. 3e).

We extended these analyses to BAU derived from RBT. In 453,163 UK Biobank participants, RBT-derived BAU was associated with mortality (HR, 1.168; 95% CI, 1.153 to 1.183), loss of healthspan (HR, 1.053; 1.044 to 1.062) and multimorbidity (HR, 1.100; 1.086 to 1.113), with positive associations across all nine incident diseases (Fig. 2g and Extended Data Fig. 3f). For external validation, we applied UK Biobank models trained on the RBT available in each cohort, retaining the UK Biobank calibration and healthy-reference scale. Higher BAU was associated with mortality in NHANES (HR, 1.211; 95% CI, 1.169 to 1.254), CHARLS (HR, 1.076; 1.020 to 1.135) and CHNS (HR, 1.227; 1.067 to 1.412; Fig. 2h).

Sensitivity analyses supported the prospective associations of BAG and BAU. In both assays, associations with all 12 outcomes remained positive after extended covariate adjustment and exclusion of events during the first year of follow-up. Association directions were also consistent across alternative follow-up periods, competing risk models and two alternative probabilistic age algorithms (Extended Data Fig. 4). Associations between higher BAU and higher risk were observed across subgroups defined by age, sex, baseline health status, BMI, smoking and physical activity, with variation in effect magnitude across participant characteristics (Extended Data Fig. 5).

Altogether, these findings support BAU as an independent indicator of future health risk beyond BAG. Associations with adverse health outcomes were consistent across NMR-derived, proteomic-derived and RBT-derived BAU, supported by subgroup analyses and replication of mortality associations in three external cohorts. Longitudinal changes in NMR-derived BAU were also associated with subsequent risk after accounting for baseline BAG and BAU. Adding BAU to conventional clinical prediction models improved risk discrimination, with models combining BAG and BAU achieving the greatest improvement.

### BAU and BAG show distinct molecular and physiological correlates

We next examined the molecular correlates of BAU using cross-omic analyses, relating NMR-derived BAU to plasma proteins and proteomic-derived BAU to NMR measures. This analysis assessed associations in a different omic layer from the features used to derive BAU, with BAG and BAU included together in each model. Higher NMR-derived BAU was associated with higher levels of the stress-responsive cytokine GDF15 and complement receptor VSIG4, and lower APOM and PON3 (Fig. 3a). Higher proteomic-derived BAU was associated with higher glucose and glycoprotein acetyls and lower albumin and free cholesterol in large LDL (Fig. 3b). These associations were replicated in held-out assessment centres, linking BAU to reproducible differences in proteins involved in stress responses and immunity, alongside glucose and lipoprotein measures.

**Fig. 3.**
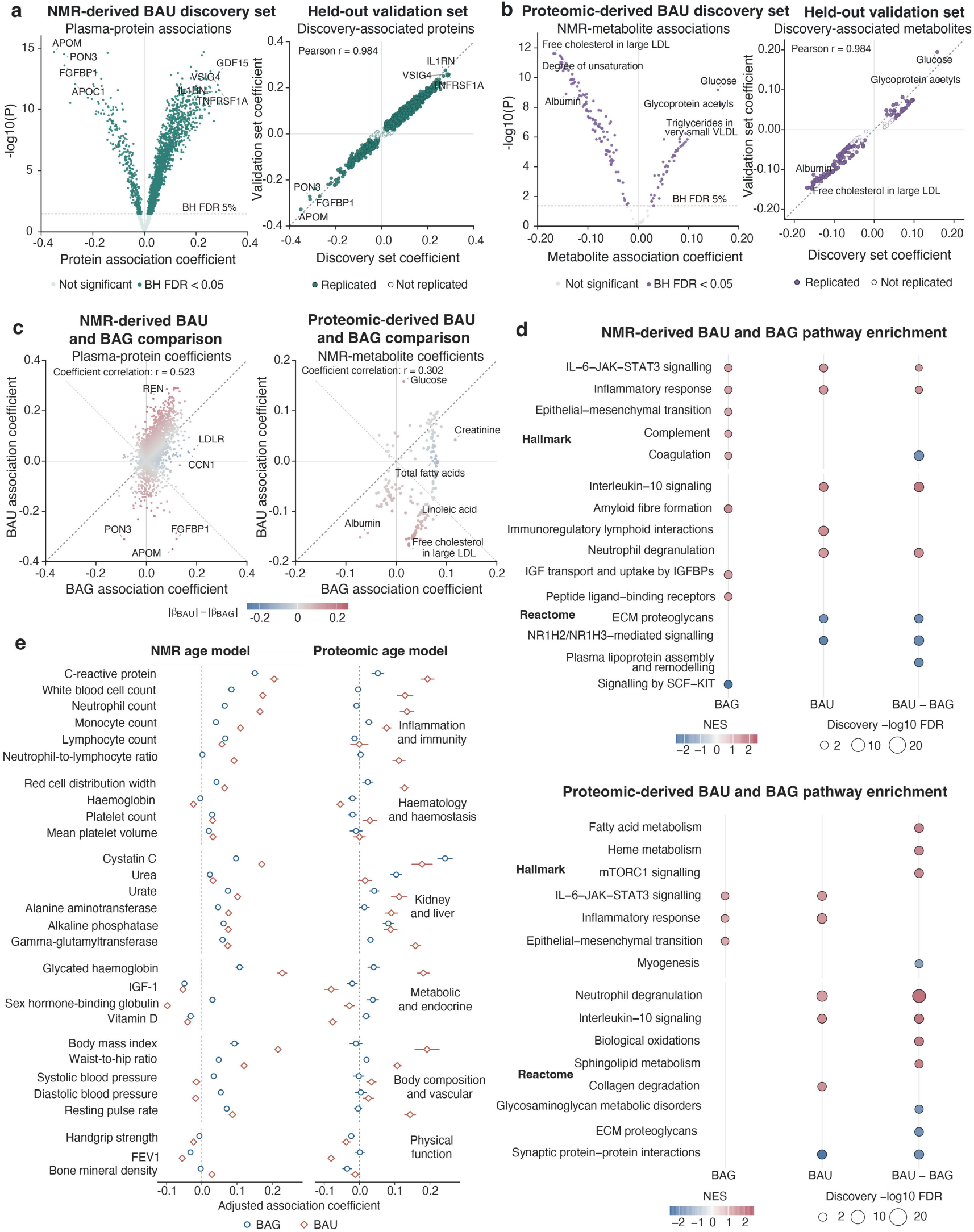
Distinct molecular, pathway and physiological correlates of BAU and BAG. **a,** Associations of NMR-derived BAU with 2,919 plasma proteins. Each point represents one protein. Left, association coefficients and statistical significance in the discovery set. Right, comparison of coefficients between discovery and validation sets from separate assessment centres, showing proteins with discovery false discovery rate (FDR) <0.05. Filled points indicate associations with the same direction and validation FDR <0.05. Models included BAG and BAU together and adjusted for age, sex, ethnicity, assessment centre, sampling date, season and molecular missingness. Coefficients represent protein SD per 1 SD higher BAU. FDR was controlled using the Benjamini-Hochberg procedure. **b,** As in **(a)**, for associations of proteomic-derived BAU with 168 NMR measures. Each point represents one NMR measure. **c,** Comparison of BAU and BAG association coefficients for plasma proteins (left) and NMR measures (right). Each point shows the BAG and BAU coefficients from the same joint regression in the discovery set. Colour indicates the difference in absolute coefficients, |βBAU| − |βBAG|. Rose indicates a larger absolute BAU coefficient, blue a larger absolute BAG coefficient and grey similar absolute coefficients. Diagonal lines mark equal absolute coefficients. Pearson correlations between BAG and BAU coefficients are indicated. **d,** Hallmark and Reactome pathway enrichment for NMR-derived BAU and BAG (top) and proteomic-derived BAU and BAG (bottom). Analyses used 2,919 plasma proteins for the NMR-derived measures and 2,715 proteins outside the proteomic age model for the proteomic-derived measures. Proteins were ranked separately by the t statistics for BAG, BAU and the difference between their association coefficients (BAU-BAG), followed by gene set enrichment analysis. Representative pathways with discovery FDR <0.05 and same-direction validation FDR <0.05 are shown. Colour indicates the normalized enrichment score, and bubble area indicates discovery -log10 FDR. **e,** Associations of BAU and BAG with 28 measured physiological traits to assess whether the molecular and pathway findings in **(a-d)** are reflected at the physiological level. Results are shown for NMR-derived BAU and BAG (left) and proteomic-derived BAU and BAG (right). Circles and diamonds show discovery coefficients for BAG and BAU, respectively; horizontal lines indicate 95% confidence intervals. Models adjusted for demographic, sampling and technical factors, together with smoking, alcohol intake, physical activity, education, income and employment. NMR, nuclear magnetic resonance.

We then compared the molecular associations of BAU with those of BAG to determine whether the two measures captured similar or distinct molecular profiles. Their association estimates were correlated across proteins (r = 0.523) and NMR measures (r = 0.302), but differed in both magnitude and direction (Fig. 3c). Glucose and albumin showed larger absolute association coefficients for proteomic-derived BAU, whereas creatinine showed a larger coefficient for proteomic-derived BAG. APOM and FGFBP1 had positive coefficients for NMR-derived BAG and negative coefficients for NMR-derived BAU. These comparisons indicate that BAU and BAG share some molecular correlates while showing distinct associations with specific proteins and metabolic measures.

To interpret these molecular differences, we compared protein pathway enrichment for BAG, BAU and their coefficient contrast. Both measures were associated with IL-6–JAK–STAT3 signalling and inflammatory response pathways, whereas interleukin-10 signalling and neutrophil degranulation showed more positive associations with BAU than with BAG. In the NMR model, NMR-derived BAU showed negative associations with NR1H2/NR1H3 signalling and extracellular matrix proteoglycan pathways, whereas NMR-derived BAG showed more positive associations with coagulation and lipoprotein remodelling. In the proteomic model, proteomic-derived BAU was associated with collagen degradation, and coefficient comparisons identified differences involving fatty acid metabolism, myogenesis and extracellular matrix pathways (Fig. 3d). Gene Ontology analyses further supported these functional differences through enrichment of immune, granule, lipoprotein and extracellular matrix processes (Extended Data Fig. 6a,b). Together, these findings indicate that BAU and BAG share inflammatory associations but differ in their associations with immune regulation, lipid metabolism and tissue remodelling.

We next assessed whether these molecular differences were reflected in independently measured physiological traits. Across 28 traits, both NMR-derived and proteomic-derived BAU showed stronger positive associations than the corresponding BAG measures with C-reactive protein, neutrophil-to-lymphocyte ratio, body mass index and waist-to-hip ratio. Proteomic-derived BAG showed stronger associations with cystatin C and urea (Fig. 3e). The associations with inflammatory and adiposity-related traits provided physiological counterparts to the molecular findings, while the renal function markers further distinguished the correlates of BAG and BAU.

We performed sensitivity analyses to assess the robustness of the molecular findings. Association patterns remained largely consistent after additional adjustment for absolute BAG, allowing nonlinear associations with BAG, excluding extreme BAG values or restricting analyses to participants with low assay missingness. Among participants healthy at baseline, molecular associations generally retained their direction, although their magnitudes were attenuated, and some pathway differences were attenuated (Extended Data Fig. 6c-e). These results support the robustness of the molecular findings across modelling choices and sample restrictions, with the primary association patterns also evident in participants healthy at baseline.

To sum up, these findings distinguish the molecular and physiological correlates of BAU from those of BAG. BAU showed prominent associations with immune, metabolic and tissue-remodelling processes, together with physiological markers of inflammation and adiposity. Differences from BAG extended across molecular association strength and direction, pathway enrichment and clinical traits.

### BAU and BAG have overlapping but distinct genetic architectures

We performed sex-stratified genome-wide association studies (GWASs) followed by cross-sex meta-analysis for the six BAG and BAU traits derived from RBT, NMR metabolomics and plasma proteomics. We identified 411 non-MHC genomic loci, defined by merging overlapping FUMA risk intervals across traits (Supplementary Tables 1-2). Manhattan and quantile-quantile (Q-Q) plots for sex-stratified GWASs, cross-sex meta-analyses and pooled-sample GWASs are shown in Fig. 4a, Extended Data Fig. 7 and Supplementary Figs. 1-6. SNP-based heritability estimates from linkage disequilibrium score regression (LDSC) supported a heritable component in all six traits (Fig. 4b). Across the six cross-sex meta-analyses, LDSC intercepts ranged from 1.000 to 1.164, and attenuation ratios ranged from 0.010 to 0.122 (Supplementary Table 3). These ratios were consistent with predominantly polygenic contributions to test-statistic inflation. In sensitivity analyses, LDSC-intercept calibration reduced inflation and significant SNP counts, while the genetic correlations, conditional associations and MAGMA gene rankings examined remained largely unchanged (Supplementary Fig. 7, Supplementary Tables 4-7, and Supplementary Note 5).

**Fig. 4.**
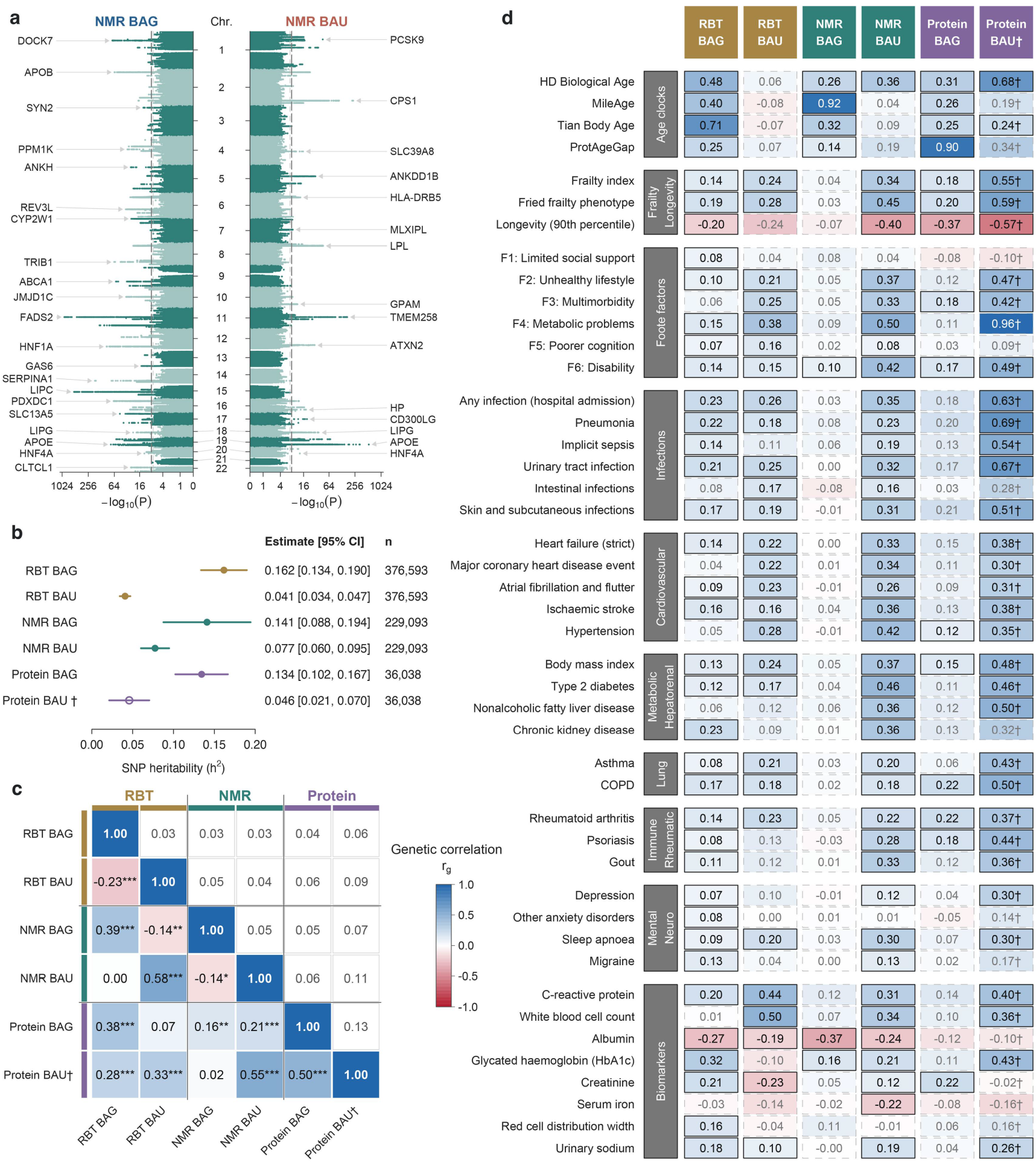
Genetic architecture and correlations of BAG and BAU. **a,** Vertical Miami plots of genome-wide association study (GWAS) results for NMR-derived BAG and BAU. Horizontal axes display log_2_[1-log_10_(P)], with tick labels showing -log_10_(P). Light dashed and dark long-dashed lines indicate P = 5×10^−8^ and P = 8.33×10^−9^ (5×10^−8^/6), respectively. Labels identify the nearest genes to the most significant variant meeting the adjusted threshold on each chromosome. GWAS tests were two-sided. **b,** Single-nucleotide polymorphism (SNP) heritability estimates obtained using linkage disequilibrium score regression (LDSC) from cross-sex GWAS meta-analyses. Points and horizontal lines indicate estimates and 95% confidence intervals (CIs), respectively. Genetic correlations involving proteomics-derived BAU were considered exploratory because of its relatively low heritability Z-score (Z = 3.66), denoted by open point. **c,** Pairwise genetic correlations among the six BAG/BAU traits estimated using joint LDSC. The lower and upper triangles show genetic correlation (*r_g_*) estimates and standard errors (SEs), respectively. Asterisks indicate significance after Benjamini-Hochberg (BH) false discovery rate (FDR) correction across 15 pairwise comparisons: q < 0.05 (*), q < 0.01 (**) and q < 0.001 (***). **d,** LDSC genetic correlations with 45 selected external phenotypes. Solid and dashed borders indicate global BH-FDR q < 0.05 and q ≥ 0.05, respectively. Correlation tests in **(c)** and **(d)** were two-sided. Blue and red indicate positive and negative correlations, respectively.

Among loci associated with RBT- and NMR-derived BAU, 54.4% (31/57) and 47.7% (21/44), respectively, overlapped with loci associated with the corresponding BAG. Shared loci, such as APOB and CETP/HERPUD1, have been linked to aging and longevity in previous studies^24,25^. Nevertheless, shared variants could have opposing associations. The T alleles of PCSK9 rs11591147 and APOE rs7412 were significantly associated with lower BAG but higher BAU in both RBT- and NMR-derived traits. Beyond these shared loci, loci containing SLC39A8, GPAM, IL1RN and ANGPTL4 reached the adjusted genome-wide significance threshold (P < 8.33 × 10^−9^) in both BAU traits but neither corresponding BAG trait. These candidate genes participate in manganese homeostasis and glycosylation^26^, triglyceride synthesis^27^, antagonism of IL-1 signalling^28^ and regulation of lipoprotein lipase^29^. Together, these findings suggest partly distinct biological underpinnings of BAG and BAU.

Genetic correlations among the six traits and with aging-related phenotypes further distinguished BAG from BAU. Genetic correlations involving proteomic-derived BAU were considered exploratory because of its relatively low heritability Z-score (Z = 3.66). RBT- and NMR-derived traits showed positive genetic correlations between their BAG measures and between their BAU measures. By contrast, BAG and BAU were negatively correlated within both RBT-derived and NMR-derived traits (Fig. 4c, Supplementary Table 8). BAG exhibited strong genetic concordance with established age-gap indicators derived from corresponding biomarker sources (Fig. 4d, Supplementary Table 9), such as NMR-derived BAG with MileAge (rg = 0.921, s.e. = 0.025), RBT-derived BAG with Tian Body Age (rg = 0.706, s.e. = 0.028) and proteomic-derived BAG with ProtAgeGap (rg = 0.904, s.e. = 0.024). Among BAU comparisons with these indicators, only proteomic-derived BAU with Tian Body Age remained significant after global FDR correction. Nevertheless, NMR- and proteomic-derived BAU correlated positively with homeostatic dysregulation (HD) Biological Age (NMR: rg = 0.355, s.e. = 0.082; proteomics: rg = 0.681, s.e. = 0.156). HD quantifies multivariate biomarker deviations from a reference distribution using Mahalanobis distance^30^. This emphasis on multivariate deviation provides a conceptual parallel to the predictive dispersion captured by BAU.

Beyond their contrasting relationships with established aging clocks, BAG and BAU also differed in their genetic correlations with aging-related phenotypes (Fig. 4d, Supplementary Fig. 8). RBT-derived BAU had higher positive correlation estimates with C-reactive protein and white blood cell count than RBT-derived BAG. The contrast was particularly pronounced for NMR-derived traits across frailty, infection and cardiometabolic outcomes. NMR-derived BAU correlated positively with Fried frailty (rg = 0.448, s.e. = 0.050) and hospitalization for infection (rg = 0.352, s.e. = 0.054). It also showed positive correlations with cardiovascular diseases (rg = 0.379, s.e. = 0.068) and type 2 diabetes (rg = 0.459, s.e. = 0.067), whereas corresponding NMR-derived BAG estimates were generally close to zero. NMR-derived BAU additionally showed a negative correlation with longevity, extending this pattern from disease-related traits to survival. Exploratory analyses of proteomic-derived BAU showed a similar correlation pattern. To assess whether these associations persisted after accounting for BAG, we fitted genetic covariance models jointly including BAG and BAU derived from the same biomarker source. Across 45 representative external phenotypes, the principal genetic associations of RBT- and NMR-derived BAU persisted after adjustment for the corresponding BAG (Extended Data Fig. 8, Supplementary Table 10). Together, these findings distinguish BAU genetically from BAG and support its potential to capture multisystem physiological dysregulation.

Sex-stratified analyses showed lower cross-sex genetic correlations for RBT- and NMR-derived BAU than for their BAG counterparts (Extended Data Fig. 9a-c, Supplementary Table 11). For external phenotypes, RBT-derived BAU showed stronger immune-inflammatory and frailty-related correlations in females. By contrast, NMR-derived BAU showed stronger correlations with selected cardiometabolic traits in males (Extended Data Fig. 9d,e; Supplementary Table 12). Further details are provided in Supplementary Note 6.

### Functional mapping reveals shared and distinct tissue and pathway enrichment patterns for BAG and BAU

To identify the tissue and cellular contexts underlying these genetic associations, MAGMA^31^ and DEPICT^32^ analyses identified significant liver and hepatocyte enrichment, respectively, across RBT- and NMR-derived BAG and BAU (Fig. 5a, Supplementary Tables 13-14). The main functional comparisons focused on RBT- and NMR-derived traits because proteomics-derived traits yielded fewer associated loci and prioritized genes, particularly BAU. Additional MAGMA enrichments included the kidney cortex and medulla, thyroid and pancreas for RBT-derived BAG, and visceral adipose tissue and spleen for RBT-derived BAU. DEPICT further implicated the hematopoietic system, myeloid cells, neutrophils and monocytes for RBT-derived BAU, and adipose tissue, monocytes and dendritic cells for NMR-derived BAU. These findings placed BAG and BAU within overlapping metabolic and hepatic tissue contexts while identifying additional immune-related enrichment for BAU.

**Fig. 5.**
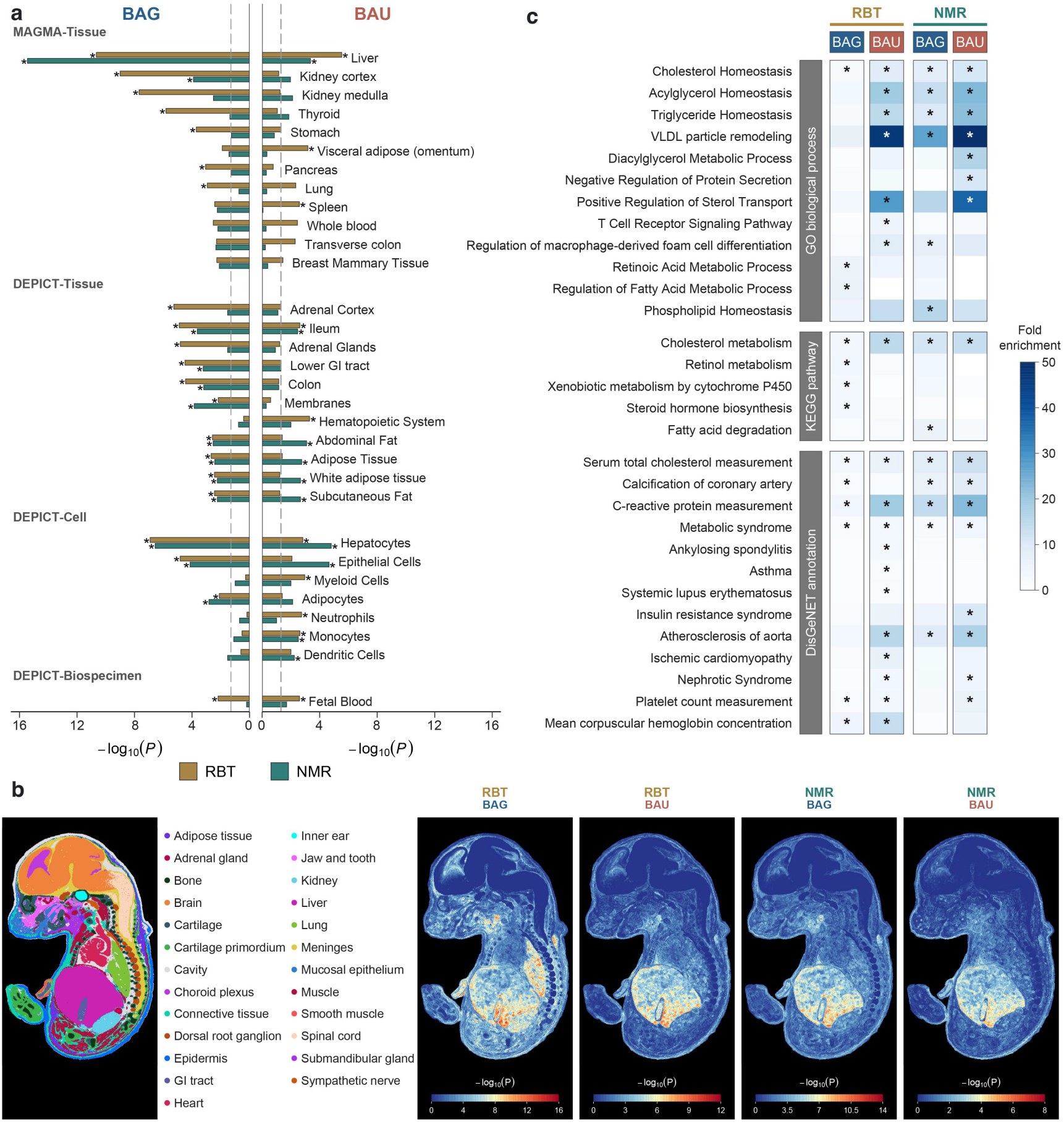
Tissue, spatial and functional mapping of BAG and BAU genetic associations. **a,** Representative MAGMA and DEPICT enrichment results for RBT-derived and NMR-derived BAG/BAU. Bars show -log_10_(P); dashed lines indicate nominal P = 0.05. Asterisks denote global Benjamini-Hochberg (BH) false discovery rate (FDR) q < 0.05 for MAGMA and DEPICT-reported within-trait FDR < 0.05. Enrichment tests were one-sided. **b,** Tissue annotations and gsMap associations in the mouse embryonic section at embryonic day 16.5 (E16.5). Colours from blue to red indicate increasing −log_10_(P) from one-sided gsMap tests. All panels use identical spatial coordinates. **c,** Selected enrichments from Gene Ontology (GO), the Kyoto Encyclopedia of Genes and Genomes (KEGG) and the DisGeNET gene-disease association database. Analyses included genes supported by at least three of five prioritization approaches. Blue intensity indicates fold enrichment relative to the database-specific gene background. Asterisks denote global BH-FDR q ≤ 0.05 from one-sided hypergeometric tests.

We next examined the spatial distribution of these associations using gsMap and E16.5 mouse embryonic Stereo-seq data from the Mouse Organogenesis Spatiotemporal Transcriptomic Atlas (MOSTA)^33^. At embryonic day 16.5 (E16.5), RBT- and NMR-derived BAG and BAU showed significant associations with the liver, gastrointestinal tract and pancreas across all sections containing these annotations (Fig. 5b, Supplementary Table 15). These tissue-level association patterns were consistent between the two E16.5 embryos (Pearson’s r = 0.864-0.942; Extended Data Fig. 10a; Supplementary Tables 16-17). Results across developmental stages are presented in Supplementary Fig. 9. To resolve the human cellular contexts of these associations, we applied scDRS^34^ to the Tabula Sapiens v2 multi-tissue scRNA-seq atlas (Extended Data Fig. 10b). All four RBT- and NMR-derived traits showed significant tissue-level associations with the liver and small intestine, alongside cell-type associations with hepatocytes and enterocytes (Supplementary Tables 18-19). Lung macrophages showed prominent signals for RBT-derived BAU and both NMR-derived traits, with the highest scores for NMR-derived BAG (Supplementary Table 20). RBT- and NMR-derived BAU also showed higher monocyte scDRS scores than their BAG counterparts, with RBT-derived BAU reaching significance after global FDR correction (Supplementary Table 19). These findings localized shared associations to hepatic and intestinal tissues while highlighting additional myeloid-cell signals for BAU.

To characterize candidate causal genes for each trait, we used five complementary gene-prioritization approaches (Extended Data Fig. 10c): positional mapping linked associated variants to nearby genes; eQTL and chromatin-interaction mapping identified regulatory connections to genes; MAGMA aggregated SNP associations into gene-level tests; and SMR-multi integrated GWAS results with expression, splicing and methylation QTLs. Requiring support from at least three approaches prioritized 1,768 and 336 genes for RBT-derived BAG and BAU, respectively, and 571 and 266 genes for NMR-derived BAG and BAU (Supplementary Table 21). We performed over-representation analysis of the prioritized genes using hypergeometric tests against Gene Ontology, KEGG and DisGeNET gene sets (Fig. 5c, Supplementary Table 22).

Cholesterol homeostasis and cholesterol metabolism were enriched across all four traits, indicating a shared lipid-metabolic background. Within this background, BAG and BAU showed different functional enrichment profiles. RBT-derived BAG showed enrichment for retinoic acid and xenobiotic metabolism, whereas NMR-derived BAG was enriched for phospholipid homeostasis and fatty acid degradation. Both BAU traits showed enrichment for positive regulation of sterol transport, with CETP, LIPG and NR1H3 contributing to this shared enrichment. Whereas CETP and LIPG were prioritized across all four traits, NR1H3 was prioritized for both BAU traits but neither BAG trait. The shared 11p11.2 locus containing NR1H3 was represented by rs35184771 in RBT-derived BAU (P = 1.54 × 10^−16^) and rs113253936 in NMR-derived BAU (P = 1.24 × 10^−10^). NR1H3 encodes LXR α, linking these findings to the established role of LXR signaling in coordinating lipid metabolism and macrophage inflammatory responses^35^. Additionally, RBT-derived BAU showed enrichment for T-cell receptor signaling. The immune-related profile of RBT-derived BAU was further supported by DisGeNET enrichment for gene sets associated with asthma and systemic lupus erythematosus. These representative enrichment patterns were retained under alternative gene-background and prioritization criteria (Supplementary Table 23). Together, these analyses highlighted differing enrichment profiles within a shared lipid-metabolic background, with additional links to immune-inflammatory processes particularly evident for BAU.

## Discussion

Our study introduces a probabilistic framework for characterizing biological age through both mean age deviation and uncertainty, quantified by BAG and BAU, respectively. Within this framework, individuals with similar BAG differed substantially in BAU, with greater BAU associated with more discordant aging, reflected in molecular age-signal disagreement and organ-age disagreement. Higher BAU was associated with mortality, loss of healthspan and multimorbidity beyond BAG, while incorporating both measures improved clinical risk discrimination. Associations with subsequent health decline were also evident for longitudinal increases in BAU, and RBT-derived BAU showed mortality associations in three independent external cohorts. Cross-omic and genetic analyses revealed distinct molecular profiles and genetic architectures for BAU and BAG, with BAU showing prominent links to immune and metabolic regulation. Collectively, these findings establish BAU as a biologically informative dimension of human aging and show how a distributional representation of biological age can connect variation in the coherence of aging signals to differences in future health vulnerability.

Our findings show that a probabilistic representation of biological age contains two distinct dimensions. Although aging clocks have progressively expanded from predicting chronological age^15^ to capturing multisystem physiological state^36,37^, mortality risk^38^, longitudinal pace of aging^39^ and organ-specific molecular changes^1,8^, each modeled aspect remains typically represented by a single scalar estimate^40^. For age prediction models, this scalar locates an individual along an aging axis, making BAG the primary phenotype of interindividual variation. In our probabilistic framework, BAG describes the deviation of mean predicted age from chronological age, whereas BAU characterizes the width of the predicted-age distribution. Individuals with similar mean predicted ages can therefore differ in the uncertainty associated with those estimates. Consistent with this distinction, participants with closely matched BAG exhibited markedly different BAU (Fig. 1c), and the two measures were weakly correlated in both the NMR and proteomic models (Fig. 1d). Considering both the mean and width of the age distribution provides a two-dimensional framework for characterizing individual differences in biological aging.

The clinical value of BAU lies in how it transforms the health-related interpretation of BAG. Previous studies have established BAG as the principal metric linking biological clocks to mortality and aging-related diseases^1,3,9^, while individual uncertainty has conventionally been treated as a measure of statistical precision or evidential strength^18,19^. Our findings demonstrate that this uncertainty arises not only from statistical uncertainty but also from underlying discordant aging process, with biological relevance, such as directly reflecting clinical vulnerability. Across both low- and high-BAG strata, individuals with high BAU exhibited elevated 15-year risks of mortality, loss of healthspan, and multimorbidity (Fig. 2a,c), maintaining independent predictive value after adjusting for BAG and established clinical risk factors (Fig. 2b,d). High BAU can thus uncover hidden susceptibility even among individuals classified as biologically young by BAG alone. Furthermore, combining BAG and BAU achieved improved risk prediction across outcomes, with longitudinal increases in BAU offering additional predictive value for future health decline. Evaluating BAU alongside BAG thus differentiates individuals with similar estimated biological age but diverging health risk, establishing both the location and uncertainty of an age distribution as essential components of biological aging assessment and risk stratification.

The associations of BAU with molecular age-signal disagreement and organ-age disagreement link uncertainty to discordant aging across biological systems. Higher NMR-derived and proteomics-derived BAU were associated with greater disagreement among molecular age signals within their respective assays (Fig. 1f-g), and higher proteomic-derived BAU was also associated with greater disagreement among organ-specific BAGs (Fig. 1h). These associations suggest that BAU reflects differences in the alignment of aging-related signals within an individual. Our mathematical framework provides a theoretical perspective for this interpretation by linking divergence among subsystem aging states to a broader predicted-age distribution and increased risk of adverse health outcomes at a given mean aging level. Discordant aging across biological systems, reflected by higher BAU, thus offers a distinct perspective on biological aging alongside the mean age deviation measured by BAG. Empirical observations across biological scales further support the biological relevance of coordinated function. Aged cardiomyocytes exhibit heightened cell-to-cell variability in gene expression^20^, and advanced age impairs coordinated transcriptional responses to immune challenge^21^. Furthermore, homeostatic dysregulation accumulates along partially independent physiological axes, predicting frailty, chronic disease, and mortality^22^, while physiological resilience depends on coordinated multi-system responses to restore homeostasis following perturbation^23^. Collectively, these theoretical and empirical insights provide a biological rationale for the predictive value of BAU.

The molecular divergence between BAU and BAG indicates that they capture distinct layers of biological aging. While BAG was predominantly linked to lipoprotein transport, tissue structure, and organ function, reflecting accumulated shifts in steady-state physiological capacity^41^, BAU was associated with interconnected components of the systemic stress response, including inflammatory activation, metabolic adjustment, and tissue remodeling. Among signals associated with BAU, GDF15 is induced by the integrated stress response during nutritional and cellular stress^42^ and contributes to the control of aging-related inflammation and metabolic homeostasis^43^, whereas higher glycoprotein acetyls provide a complementary signal of persistent inflammation and neutrophil activity^44^. Experimental evidence directly links these pathways, as NLRP3-mediated sterile inflammation impairs metabolic control during aging^45^, and senescent cell secretomes concurrently release GDF15 alongside matrix-remodeling proteases^46^. The simultaneous enrichment of these pathways in high BAU profiles depicts a systemic state in which stress sensing, metabolic adaptation, and tissue repair are chronically engaged. Differential activation of these pathways across biological systems may contribute to discordant aging, reflected in disagreement among age-informative signals, consistent with the progressive homeostatic dysregulation and heightened biological heterogeneity characteristic of aging^22,47^. In this view, BAG quantifies the magnitude of accumulated molecular and functional change, whereas BAU captures discordant aging across biological systems related to systemic regulatory instability.

Genetic analyses extend these molecular findings by revealing distinct health-related associations and functional enrichment profiles for BAG and BAU. Within RBT- and NMR-derived traits, negative BAG-BAU genetic correlations and opposing associations at shared variants argue against treating the two phenotypes as interchangeable measures of aging. BAU showed higher positive genetic correlation estimates than BAG for aging-related phenotypes related to inflammation, frailty and metabolic dysfunction. Key associations persisted after adjustment for BAG, supporting health-related genetic information not fully captured by BAG. The positive genetic correlation of NMR-derived BAU with HD Biological Age^30^ is consistent with BAU capturing aspects of physiological dysregulation and suggests that the uncertainty it quantifies may have biological relevance in aging-related biomarker patterns. Functional mapping places these differences within metabolic systems shared by both phenotypes. Common lipid-pathway enrichment can involve different genes and cellular contexts, allowing BAG and BAU to reflect different aspects of interconnected biological processes. The additional immune-related enrichment observed for BAU provides a functional context for its genetic associations with inflammation and frailty. The prioritization of NR1H3, encoding LXRα, in both BAU traits offers a relevant connection to established biology. LXRs coordinate cholesterol handling and macrophage inflammatory responses, providing a framework for interpreting the sterol transport and myeloid-cell associations^35^. Overall, these findings support BAU as a new dimension of aging assessment, distinct from but complementary to BAG.

This study has several limitations. First, BAU was parameterized as the log predictive variance under a Gaussian distribution, whereas alternative or non-parametric formulations may capture asymmetric or heavy-tailed uncertainty differently. Nevertheless, the strong associations observed across health outcomes, cross-omic profiles, and genetic architectures demonstrate that even a Gaussian-based representation captures biologically informative signals beyond BAG, indicating that the core concept of BAU is robust to distributional assumptions. Second, as a model-derived metric, BAU inevitably retains residual technical and modeling noise alongside biologically meaningful variation. However, measured technical and other factors accounted for only a modest proportion of BAU variance, and individual BAU rankings remained highly consistent following rigorous adjustment and across alternative model specifications. Third, the primary NMR and proteomic models were trained and evaluated within the UK Biobank, requiring further testing in external cohorts with harmonized omics depth. Nonetheless, RBT-derived BAU remained independently predictive of mortality across three external validation cohorts without cohort-specific re-fitting or recalibration, supporting the broader generalizability of this framework across diverse populations.

This study also has several strengths. First, to our knowledge, this work provides the first population-scale characterization of BAU as an essential dimension of human aging, spanning longitudinal health outcomes, multi-omic profiling, and genome-wide genetic analyses. By jointly modeling central tendency and dispersion, our probabilistic framework moves beyond single scalar age estimates to represent biological age as a distribution, simultaneously quantifying BAG and BAU. Second, BAU demonstrated clear clinical utility by providing predictive information beyond BAG across mortality, healthspan loss, multimorbidity, and diverse incident diseases. Importantly, both baseline BAU and its longitudinal change independently tracked future health decline, while RBT-derived BAU successfully generalized across independent populations, highlighting its potential for individualized risk stratification and clinical monitoring. Third, integrated multi-omic and genetic analyses established distinct biological foundations for BAU and BAG, disentangling the biology of BAU from that of BAG. Together, cross-omic signal divergence alongside specific immune, metabolic, and tissue-remodeling signatures establishes discordant aging as a hallmark of high BAU.

In conclusion, our findings redefine biological age as a probabilistic state characterized jointly by its position and its uncertainty. BAG quantifies the position of an inferred age state from chronological age, whereas BAU measures how broadly biological signals disperse across compatible age states. Comprehensive evidence from prospective health outcomes, multi-omic profiles, and genetic analyses establishes BAU as an independent and essential dimension of human biological aging beyond BAG. Individual variation in aging thus spans two distinct axes, the position of an age state and the degree of discordant aging across biological systems. A distributional representation of biological age therefore provides a comprehensive framework for understanding aging heterogeneity and evaluating individual vulnerability to future health decline.

## Methods

### Ethics

UK Biobank has Research Tissue Bank approval from the North West Multi-centre Research Ethics Committee (21/NW/0157). All participants provided written electronic informed consent. NHANES protocols were approved by the National Center for Health Statistics Ethics Review Board (98-12, 2005-06, 2011-17 and 2018-01). CHARLS was approved by the Biomedical Ethics Review Committee of Peking University (IRB00001052-11014 and IRB00001052-11015). CHNS was approved by the institutional review boards of the University of North Carolina at Chapel Hill and the National Institute of Nutrition and Food Safety, Chinese Center for Disease Control and Prevention, and by the Human and Clinical Research Ethics Committee of the China-Japan Friendship Hospital. Participants in NHANES, CHARLS and CHNS provided written informed consent.

### Study populations

The primary analyses were conducted in UK Biobank, a prospective cohort of approximately 500,000 participants recruited at 22 assessment centres across England, Scotland and Wales between 2006 and 2010. Participants were aged 40 to 70 years at recruitment and provided questionnaire data, physical measurements and biological samples.

Chronological age at assessment was calculated from the attendance date and recorded month and year of birth and expressed to one decimal place. We included participants aged 40 to 70 years with available data on age, sex, assessment centre and baseline disease history. The NMR, proteomic and RBT populations were defined separately to maximize the available sample for each measurement platform. After assay-specific eligibility and missingness criteria were applied, the analysis populations comprised 272,672 participants for NMR, 43,867 for proteomics and 453,163 for RBT (Supplementary Table 24). Longitudinal analyses included 13,620 participants with NMR measurements at both baseline and reassessment (Supplementary Table 25).

External mortality analyses included participants from the US National Health and Nutrition Examination Survey (NHANES; 1999-2018), the 2011 baseline wave of the China Health and Retirement Longitudinal Study (CHARLS), and the 2009 biomarker wave of the China Health and Nutrition Survey (CHNS). We restricted these analyses to participants aged 40 to 70 years with the blood measurements required to harmonize the routine blood tests (RBT) predictors with those used in UK Biobank. Baseline characteristics of these external cohorts are summarized in Supplementary Table 26. Analysis-specific sample sizes, outcome events and follow-up are summarized in Supplementary Table 27.

### Molecular and clinical measurements

UK Biobank plasma metabolites were quantified by Nightingale Health using proton nuclear magnetic resonance (NMR) spectroscopy. Percentage and ratio variables were excluded from the original panel of 249 measures, leaving 168 absolute measures for analysis^48^. Plasma proteins were measured using the Olink Explore platform. The proteomic age model used 204 proteins selected from a previously published panel^15^. Molecular association analyses examined 2,919 proteins that met the missingness criteria. Protein association and pathway analyses of proteomic-derived biological age gap (BAG) and biological age uncertainty (BAU) excluded the 204 age-model inputs, leaving 2,715 proteins. The RBT age model included 31 haematological and 28 serum biochemistry measurements^2^, with sex as the only non-blood predictor^1^. Full feature lists and corresponding UK Biobank field identifiers are provided in Supplementary Table 28.

### Baseline health and covariates

The healthy reference population comprised participants without a baseline history of 23 chronic conditions spanning cardiovascular, respiratory, metabolic, renal, neurological, psychiatric, inflammatory, hepatic, musculoskeletal and malignant diseases (Supplementary Table 29). Healthy participants formed the model-development population. Within each fold, separate subsets were used for model fitting and calibration, with the calibration subset also providing the age and sex reference functions and standardization parameters for BAG and BAU. Unless otherwise stated, downstream analyses included all eligible participants irrespective of baseline health status.

Primary prospective analyses adjusted for chronological age, sex, current smoking, alcohol intake, moderate-to-vigorous physical activity, education, household income and employment status. Molecular association analyses adjusted for age, sex, ethnicity, assessment centre, assessment date, season and missingness in the molecular outcome assay. Physiological analyses additionally included behavioural and socioeconomic covariates and accounted for missingness in the age-model inputs. Covariate definitions and coding are provided in Supplementary Tables 30-31.

### Mathematical representation of biological aging and aging heterogeneity

#### Aging representation

Consider an individual *j* whose biological system consists of *m* heterogeneous subsystem aging signals denoted as *X_ji_*.

Assume that subsystem aging signals contribute to overall aging through a common smooth nonlinear damage function ℎ(⋅), such that the latent overall aging is

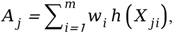

where *w_i_ > 0* denotes subsystem-specific contribution to overall aging, with 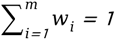.

#### Decomposition of overall aging

Define the weighted mean and weighted variance of subsystem as:

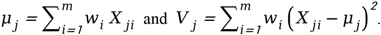

We have

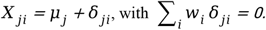

### A second-order Taylor expansion of ℎ(*X_ji_*) around *μ_j_* gives

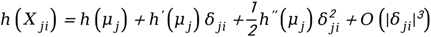

where

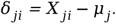

Substituting this expansion into

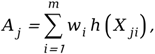

yields

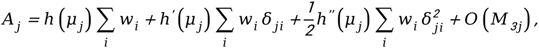

Where

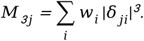

We have

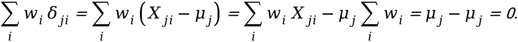

Furthermore,

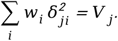

Therefore,

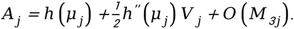

Thus, to second order, overall aging *A_j_* is characterized by two macroscopic quantities: the weighted mean aging state *μ_j_*, representing the weighted mean of subsystem aging signals, and the weighted variance *V_j_*, representing heterogeneity in subsystem aging states, a theoretical representation of discordant aging.

#### Association with overall aging

We assume ℎ′(*x*) *>0* because biological damage increases monotonically with aging, and ℎ′′(*x*) *>0* because aging-related damage accumulates at an accelerating rate (Supplementary Note 1).

From the second-order decomposition above,

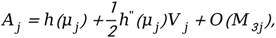

where *O*(*M_3j_*) collects third- and higher-order contributions arising from the shape of the subsystem-aging distribution. The weighted mean *μ_j_* therefore determines the leading-order level of overall aging. Moreover, conditional on *μ_j_*, the second-order contribution of subsystem heterogeneity is

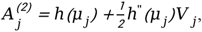

so that

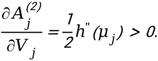

Thus, within the second-order approximation, greater subsystem aging heterogeneity increases overall aging even at the same weighted mean aging level. The same second-order argument implies that greater subsystem heterogeneity is associated with higher risk of mortality and other aging-related outcomes when these outcomes increase monotonically with overall aging (Supplementary Note 2).

#### Identification using NGBoost

To estimate *μ_j_* and *V_j_* from *X_j_*, we instead assume that the conditional distribution of chronological age *Y_j_* given *X_j_* identifies monotone transformations of these two latent quantities: *E(Y_j_*|*X_j_)=f(μ_j_)* and Var (*Y_j_*|*X_j_*) *=q* (*V_j_*)), where *f* and *q* are strictly increasing. Supplementary material gives a sufficient generative model under which these relations arise exactly (Supplementary Note 3). Using NGBoost with a heteroscedastic Gaussian conditional model, we have

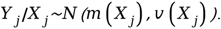

The fitted distribution provides individual-level estimates m̂_j_ and v̂_j_ of the conditional mean E(Y_j_ | X_j_) and variance Var(Y_j_ | X_j_), respectively. These estimates serve as observable proxies for latent aging level and aging heterogeneity. Under the monotonic identification assumption, the conditional mean and variance preserve the ordering of latent aging level and heterogeneity, respectively (Supplementary Note 4).

### Probabilistic age models

Separate probabilistic age models were developed for the NMR, proteomic and RBT panels, with sex included as an additional predictor^1^. Participants missing at least 20% of measurements in the relevant panel were excluded, as were features with at least 20% missingness. Remaining missing predictor values were imputed using medians estimated in the corresponding model-fitting set.

Assessment centres were assigned to five outer folds. In each iteration, all eligible participants from the held-out centres formed the test set, and healthy participants from the remaining centres formed the development sample. The development sample was divided into an 80% model-fitting set and an independent 20% healthy calibration set. Imputation parameters, preprocessing transformations, hyperparameters and model parameters were estimated in the model-fitting set. Predictions from the five held-out test sets were pooled to obtain one out-of-fold predictive distribution per participant.

The primary learner was natural-gradient boosting (NGBoost)^49^ with a Gaussian conditional distribution for chronological age. Given the molecular or RBT and sex encoded in the predictor vector **x**_i_ for participant i, the model specified:

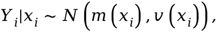

where *Y_i_* denotes chronological age, and *m* (*x_i_*) and *v* (*x_i_*) denote the conditional mean and variance, respectively. The fitted model provided individual-level estimates

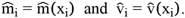

Hyperparameters were selected within the model-fitting set by 5-fold cross-validation minimizing Gaussian negative log likelihood. Gaussian location-scale models implemented using LightGBMLSS^50^ and XGBoostLSS^51^ were used as alternative probabilistic learners in sensitivity analyses.

### Derivation of BAG and BAU

Predictive means and variances were calibrated separately by sex using the independent healthy calibration set within each outer fold. For each sex s, chronological age was regressed on the predicted mean using an affine calibration model, yielding the calibrated predictive mean:

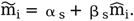

Predictive variances were then rescaled using prediction errors obtained from cross-fitted mean calibration within the same healthy calibration set:

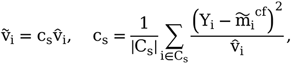

where C_s_ denotes the sex-specific healthy calibration set and m̃_i_^cf^ denotes the corresponding cross-fitted calibrated mean. This rescaling aligned the magnitude of the predicted variances with the observed squared prediction errors in the healthy calibration set.

Raw biological age gap and uncertainty were defined as:

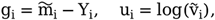

where log denotes the natural logarithm.

To remove residual age dependence, we fitted sex-specific LOWESS curves for calibrated age gap and log predictive variance as functions of chronological age in the healthy calibration set. BAG and BAU were calculated by subtracting the corresponding fitted age trends and dividing by the residual standard deviations in that set:

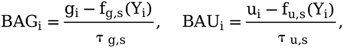

where τ_g,s_ and τ_u,s_ denote the sex-specific residual standard deviations in the healthy calibration set. Positive BAG therefore indicates a higher mean predicted age than expected for a healthy participant of the same chronological age and sex, whereas positive BAU indicates greater uncertainty than expected on the same reference scale. Calibration and reference parameters were estimated in each fold’s healthy calibration set and applied unchanged to participants in the held-out centres.

### Evaluation of BAU

Age prediction performance was evaluated using calibrated mean predicted ages from held-out assessment centres. Pearson correlations with chronological age and mean absolute errors were calculated for the pooled predictions and separately within each outer fold. To examine the relationship between BAG and BAU, we calculated their Pearson correlation separately for NMR-derived and proteomic-derived measures. Illustrative predicted-age distributions were selected from NMR participants of the same age and sex, matching calibrated mean predicted age within negative and positive BAG ranges to compare participants with similar BAG but different BAU.

Longitudinal consistency was assessed using paired baseline and repeat NMR measurements. BAG and BAU at reassessment were calculated using the same age models, preprocessing parameters, calibration functions and healthy-reference parameters used at baseline.

Spearman correlations between baseline and reassessment values were calculated for both measures to compare their consistency over time.

To assess how much BAU variation was explained by potential confounders, we fitted auxiliary LightGBM regression models with BAU as the outcome. Models were fitted in four assessment-centre folds and evaluated in the fifth, yielding one out-of-fold prediction per participant. The reference model included chronological age, sex and BAG. Four groups of predictors were added separately to this model, covering technical factors, lifestyle and socioeconomic factors, medication use and baseline clinical characteristics (Supplementary Tables 30-31). A further model included all four groups together. Total R² was calculated from the pooled out-of-fold predictions for each model, with 95% confidence intervals obtained from 2,000 bootstrap resamples of assessment centres using the fixed predictions. Adjusted BAU was calculated by subtracting each participant’s out-of-fold prediction from the all-factor model from their observed BAU. We then calculated the Spearman correlation between original and adjusted BAU, with 95% confidence intervals estimated by leave-one-centre-out jackknife.

Consistency across algorithms was assessed by comparing BAU estimates from NGBoost, LightGBMLSS and XGBoostLSS. The algorithms used the same assessment-centre folds, healthy model-fitting and independent calibration sets, and healthy-reference standardization procedure. Pairwise Spearman correlations were calculated in the same participants, separately for NMR-derived and proteomic-derived BAU.

### Molecular and organ-age disagreement

We examined whether higher BAU, which indicates a broader predicted-age distribution, was associated with greater disagreement among age signals within an individual. At the molecular level, we assessed how well a common age could describe a participant’s protein or NMR measurements. At the organ level, we quantified differences among organ-specific BAGs. We then examined the associations of these disagreement measures with BAU after adjustment for overall BAG.

Molecular disagreement was calculated separately using the 204 protein inputs and 168 NMR measures of the corresponding age models. For each molecular measure, we first estimated how its expected level varied with age in healthy participants. Within each outer fold, sex-specific reference curves were fitted using natural cubic splines of age with four degrees of freedom in the healthy model-fitting set. The independent healthy calibration set was then used to correct the mean residual and estimate the residual standard deviation. These reference curves and residual standard deviations were used to calculate the likelihood of each held-out participant’s observed measurements across candidate ages from 40 to 70 years at 0.25-year intervals.

For participant i, we calculated the Gaussian log-likelihood of each observed molecular measurement x_ij_ at candidate age a as

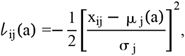

where j indexes molecular measures, μ_j_(a) is the expected level of measure j at age a after calibration, and σ_j_ is its residual standard deviation in the healthy calibration set. Both quantities were estimated separately by sex and outer fold. Age-independent constants were omitted because they cancel when calculating molecular disagreement.

For each participant, we identified the age that maximized the log-likelihood of each molecular measurement separately. We also identified a common support age 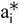 that maximized the mean log-likelihood across all observed measurements. Molecular disagreement was defined as the mean reduction in log-likelihood when each measurement was evaluated at this common support age rather than its individually best-fitting age,

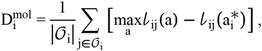

where 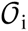 contains the indices of the molecular measures observed for participant i, and 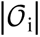 is their number. Larger values indicate that a common support age provides a poorer fit relative to allowing each molecular measure its own best-fitting age, reflecting greater disagreement among molecular age signals. The calculation used only observed measurements.

Organ-age disagreement quantified the variation in organ-specific BAGs within each participant. We constructed ten proteomic organ clocks using ridge regression and predefined protein panels (Supplementary Table 32). The clocks used the same assessment-centre folds and the same healthy participants for model fitting and calibration as the overall proteomic age model. Missing protein measurements were imputed using medians from the model-fitting set, and protein levels were standardized using means and standard deviations from that set. The ridge penalty was selected by internal cross-validation using the one-standard-error rule.

Within each outer fold, predicted organ ages were calibrated by regressing chronological age on predicted age in the healthy calibration set, separately by sex. For each organ, we then calculated the difference between calibrated predicted age and chronological age. Its association with chronological age was fitted using a natural cubic spline with four degrees of freedom in the same calibration set. The fitted age-dependent mean was subtracted from each participant’s organ age gap, and the result was divided by the calibration residual standard deviation to obtain organ-specific BAG. For participants with all ten organ BAGs available, organ-age disagreement was defined as

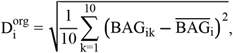

where BAG_ik_ is the standardized BAG for organ k in participant i, and 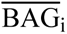 is that participant’s mean BAG across the ten organs. Larger values indicate greater differences in BAG among organs. Prediction performance was evaluated separately for each organ clock using all participants with available predictions from held-out centres. The disagreement analysis included 35,059 participants with estimates for all ten organs.

We examined molecular disagreement in relation to BAU from the corresponding assay, and organ-age disagreement in relation to overall proteomic-derived BAU. Linear regression models included BAU as the outcome and natural cubic splines with four degrees of freedom for disagreement, overall BAG and chronological age. Additional covariates were sex, assessment centre and the proportion of missing molecular measurements among the corresponding overall age model’s inputs. The organ analysis also adjusted for the proportion of missing proteins across the combined organ panels.

Adjusted curves were obtained by averaging predicted BAU across participants at each disagreement value, while retaining their observed values for the other covariates. Models included all eligible participants, and curves were displayed between the 1st and 99^th^ percentiles of disagreement. Pointwise 95% confidence intervals were calculated using robust standard errors clustered by assessment centre.

### Health outcomes and prospective analyses

UK Biobank outcomes were ascertained from self-reported information and linked primary care, hospital, cancer and mortality records. Follow-up began at baseline and ended at the target event, death, loss to follow-up or 31 December 2022, whichever occurred first. All-cause mortality was obtained from national death registry records. Loss of healthspan was defined as the first occurrence of myocardial infarction, heart failure, stroke, chronic obstructive pulmonary disease, diabetes, dementia, malignant cancer excluding non-melanoma skin cancer and carcinoma in situ, or death^52,53^. Participants with any of these diseases at baseline were excluded from this analysis. Major disease multimorbidity was defined as the occurrence of a second distinct disease domain among the same seven diseases^54^, with participants having zero or one domain at baseline included. We also examined nine incident diseases individually, comprising coronary heart disease, stroke, chronic obstructive pulmonary disease, liver disease, diabetes mellitus, renal failure, dementia, digestive system cancer and respiratory system cancer. Each analysis excluded participants with the corresponding disease at or before baseline. Detailed definitions and code lists are provided in Supplementary Table 33.

For risk curves, participants were classified into four groups according to whether BAG and BAU were below or at least zero on the healthy-reference scale. Curves for mortality, loss of healthspan and the nine individual diseases were estimated as one minus the Kaplan-Meier survival function, with death treated as censoring for individual diseases. Multimorbidity curves were estimated using the Aalen-Johansen cumulative-incidence function, treating death as a competing event. Curves were displayed through 15 years with 95% confidence intervals.

Prospective associations were estimated using Cox proportional hazards models containing BAG and BAU simultaneously. Models adjusted for chronological age, sex, current smoking, alcohol intake, moderate-to-vigorous physical activity, education, household income and employment status. Baseline hazards were stratified by assessment centre, with standard errors clustered by centre. Multimorbidity models were additionally stratified by the number of disease domains present at baseline. Death before an individual disease or multimorbidity event was treated as censoring. Hazard ratios were reported per 1 SD increase in BAG or BAU on the healthy-reference scale.

To assess whether BAG and BAU improved clinical risk prediction, we compared a baseline clinical model with models additionally containing BAG, BAU or both. The baseline model included age, sex, current smoking, diabetes history, systolic blood pressure, total cholesterol, HDL cholesterol and antihypertensive treatment. Separate Cox models were fitted for each assay and outcome using five assessment-centre folds. Each model was fitted in four folds and used to predict 10-year risk in the remaining fold. Predictions from all held-out folds were combined to calculate the Uno C-index at 10 years. We reported both the C-index for each model and its change relative to the baseline clinical model, comparing models in the same participants. Confidence intervals accounted for assessment-centre clustering and the paired predictions. These analyses covered the three primary outcomes and all nine incident diseases in the NMR and proteomic populations.

### Longitudinal analyses of BAG and BAU

We examined whether changes in NMR-derived BAG and BAU were associated with subsequent health outcomes. Changes were calculated as reassessment minus baseline values and standardized separately among the 13,620 participants with paired measurements.

Follow-up began at reassessment, with each analysis restricted to participants who remained under observation and free of the corresponding outcome. Cox proportional hazards models included both baseline measures and both changes, with additional adjustment for age at reassessment, sex, the interval between assessments and the baseline behavioural and socioeconomic covariates described above. Baseline hazards were stratified by baseline assessment centre. Outcome definitions and censoring followed those described for the baseline analyses. Hazard ratios were reported per 1 SD increase in BAG or BAU change, with 95% confidence intervals calculated using model-based standard errors.

### RBT models and external validation

To examine whether BAU derived from routinely available blood measurements was associated with future health risk, we developed a probabilistic age model in UK Biobank. Model development followed the same five centre-based outer folds, healthy-reference selection, calibration and BAG and BAU derivation procedures as the NMR and proteomic models. Associations of RBT-derived BAG and BAU with the three primary outcomes and nine incident diseases were evaluated using the prospective survival models described above.

For external validation, we trained separate models in healthy UK Biobank participants using the blood measurements available in each external cohort. The panels comprised 24 measurements for NHANES, 16 for CHARLS and 18 for CHNS (Supplementary Table 34). Preprocessing parameters and fitted age models were estimated in UK Biobank, with calibration based on an independent healthy calibration set. These parameters and models were applied unchanged to the corresponding external cohort. BAG and BAU were calculated using the age- and sex-specific healthy references established in UK Biobank.

Mortality analyses included 24,225 participants in NHANES, 9,604 in CHARLS and 5,137 in CHNS. Cox proportional hazards models included BAG and BAU simultaneously and adjusted for the demographic, behavioural and health covariates specified for each cohort (Supplementary Table 30). Hazard ratios were reported per 1 SD increase on the corresponding UK Biobank healthy-reference scale. NHANES mortality was obtained through linkage to the National Death Index. For CHARLS and CHNS, deaths recorded between survey waves were assigned to the midpoint of the corresponding interval. Sensitivity analyses used the earliest and latest plausible death dates within each interval (Supplementary Table 35).

### Sensitivity and subgroup analyses

We performed separate sensitivity analyses to assess the robustness of the prospective associations of BAG and BAU. Extended adjustment added diet, medication use, family history, body mass index, systolic blood pressure and baseline disease burden to the primary models. A one-year landmark analysis began follow-up one year after baseline among participants who remained alive, under observation and free of the target outcome. To examine associations across follow-up periods, we fitted separate models for 0-5, 5-10 and more than 10 years, including participants still at risk at the start of each interval. Fine-Gray models accounted for death as a competing event for multimorbidity and the nine nonfatal disease outcomes. We also repeated the outcome analyses using BAG and BAU derived from LightGBMLSS and XGBoostLSS, retaining the same outcome definitions and covariate adjustment.

We examined whether associations with the three primary outcomes differed by age, sex, baseline health status, body mass index, current smoking and physical activity, separately for NMR-derived and proteomic-derived measures. Each Cox model included BAG, BAU, the subgroup variable and its interactions with both BAG and BAU, with adjustment following the primary analyses. Subgroup-specific hazard ratios were estimated from these models. Differences between subgroups were assessed using global Wald tests of the interaction terms for BAG and BAU separately, with Benjamini-Hochberg correction across the interaction tests.

### Molecular association and pathway analyses

We examined whether BAU and BAG were associated with different molecular profiles. For NMR-derived BAG and BAU, we analysed associations with 2,919 plasma proteins. For proteomic-derived BAG and BAU, we analysed associations with 168 NMR measures.

Assessment-centre folds 1-3 formed the discovery set, and folds 4-5 formed the validation set. Molecular measurements below the 0.5th or above the 99.5th percentile were set to the corresponding percentile values, then standardized using means and standard deviations from healthy participants in the discovery set. All preprocessing parameters were determined in the discovery set and applied unchanged to the validation set. Each protein or NMR measure was analysed among participants with an observed value for that measure.

For each molecular outcome, we fitted a linear regression model containing both BAG and BAU to estimate their mutually adjusted associations. Models also included chronological age, sex, ethnicity, assessment centre, attendance date, season and the proportion of missing measurements in the outcome assay. Chronological age was modelled using a natural cubic spline with four degrees of freedom, and standard errors were clustered by assessment centre. Associations were tested two-sided in the discovery set, with the FDR controlled at 5%. Associations meeting this threshold were tested one-sided in the validation set in the direction observed in the discovery set. Replication required the same association direction and an FDR below 5% in the validation set.

To compare the molecular associations of BAG and BAU, we plotted their coefficients estimated in the discovery set from the same joint model for all proteins and NMR measures (Fig. 3c). Coefficient signs indicated association direction, and the continuous colour scale represented the difference in absolute coefficient magnitude, M = |β_BAU_| − |β_BAG_|. We also formally tested the signed coefficient difference, Δ = β_BAU_ − β_BAG_, using a standard error that accounted for the covariance between the two estimates. These differences were evaluated using the same criteria in the discovery and validation sets as the individual associations.

To interpret the biological processes associated with BAG and BAU, we performed pathway analyses using their protein associations. For the NMR age model, we used associations with all 2,919 proteins. For the proteomic age model, we analysed associations with the 2,715 proteins remaining after exclusion of the 204 model inputs, using the same joint regression framework. Proteins were ranked separately for BAG, BAU and the signed difference between their coefficients. Each ranking statistic was calculated by dividing the coefficient, or coefficient difference, by its standard error. Gene-set enrichment analysis used the complete ranked protein lists and the Hallmark and Reactome collections from MSigDB 2026.1.Hs. Gene sets containing 10-500 measured proteins were retained. In the discovery set, FDR was controlled separately for each ranking and collection. Pathways meeting an FDR below 5% in the discovery set were evaluated in the validation set, with FDR controlled within the corresponding selected pathway set. Replication required enrichment in the same direction and an FDR below 5% in the validation set.

We also used Gene Ontology over-representation analysis to identify functions enriched among proteins with replicated associations. Separate protein sets were defined by positive and negative associations with BAG, positive and negative associations with BAU, and positive and negative BAU-minus-BAG coefficient differences. Each set was examined for enrichment in biological processes, cellular components and molecular functions, using the eligible measured proteins mapped to each category as the background. Enrichment was assessed using one-sided hypergeometric tests, with Benjamini–Hochberg FDR correction applied separately within each protein set and Gene Ontology category.

Sensitivity analyses assessed whether the molecular findings were robust to alternative ways of accounting for BAG, restrictions on assay missingness and baseline health status. We repeated the analyses after separately adding absolute BAG to the model, modelling BAG using a natural cubic spline, restricting participants to the central 95% of BAG, restricting participants to those with low missingness in the outcome assay, or restricting participants to those healthy at baseline. BAU association coefficients were compared with those from the primary analysis. Pathway analyses were also repeated among participants healthy at baseline, using the same enrichment and validation procedures.

### Analyses of physiological and clinical traits

To examine whether the molecular differences between BAG and BAU were reflected at the physiological level, we analysed their associations with 28 physiological and clinical traits (Supplementary Table 36). Traits directly overlapping with NMR age-model inputs were excluded. The discovery and validation sets followed the same assessment-centre folds as the molecular analyses. Trait measurements were winsorized, transformed where appropriate and standardized relative to healthy participants in the discovery set. The same preprocessing parameters were applied to the validation set.

Each trait was analysed using a linear regression model containing BAG and BAU simultaneously. Models adjusted for chronological age using a natural cubic spline with four degrees of freedom, sex, ethnicity, current smoking, alcohol intake, moderate-to-vigorous physical activity, education, household income, employment status, assessment centre, attendance date, season and the proportion of missing measurements among the corresponding age-model inputs. Standard errors were clustered by assessment centre. Differences between BAG and BAU coefficients were tested while accounting for their covariance within the same model. Associations and coefficient differences were evaluated using the same discovery and validation criteria as the molecular analyses.

### Genome-wide association analyses

We analyzed BAG and BAU derived from RBT, NMR metabolomics and plasma proteomics. Calibrated, standardized phenotypes were linked to genetic covariates by participant identifier. Analyses included participants of European ancestry whose recorded sex matched their genetically inferred sex and who had complete data for all covariates. The analysis samples comprised 173,318 males and 203,275 females for RBT-derived traits, 105,521 males and 123,572 females for NMR-derived traits, and 16,571 males and 19,467 females for proteomics-derived traits, respectively. Participants could contribute to more than one biomarker source. GWASs used the calibrated, age-adjusted and standardized BAG and BAU measures described above. No additional standardization or rank-based inverse-normal transformation was applied during GWAS preparation.

Sex-stratified GWASs used REGENIE (v4.1.2) in quantitative-trait mode^55^. Step 1 fitted whole-genome predictions using the predefined array-genotype set, and Step 2 tested imputed variants on chromosomes 1-22 using leave-one-chromosome-out predictions. Block sizes were 3,000 and 1,500 variants for Steps 1 and 2, respectively. Step 2 used reference-first allele ordering and a minimum minor-allele count of 20. Both steps adjusted for age, age squared, genotyping batch, assessment centre and the first ten genetic principal components. Pooled-sample GWASs used the same framework with an additional sex covariate. These pooled analyses were reported separately from the cross-sex meta-analyses, which supplied the primary downstream summary statistics.

### Cross-sex meta-analysis and summary statistic quality control

Male and female GWASs were combined by fixed-effect inverse-variance meta-analysis using the STDERR scheme in METAL (release 5 May 2020), with genomic control disabled^56^. Between-sex heterogeneity statistics were also calculated. Variants were matched by chromosome, position and ordered allele pair. Input preparation excluded records with missing required fields or nonnumeric or nonpositive standard errors (SEs). Per-variant sample sizes were summed across contributing sexes, and meta-analysis INFO was the minimum available imputation INFO score across the contributing sex-specific files. Variants used for GWAS plots and gene mapping required INFO ≥ 0.8 and minor-allele frequency (MAF) ≥ 0.01. Extreme probabilities were recovered using stable normal-tail calculations and bounded below at 10^−300^ for FUMA uploads; extended -log_10_(P) values were retained for plotting. FUMA inputs were restricted to P ≤ 0.05, whereas MAGMA and SMR used the full eligible summary statistics. The adjusted significance threshold was P ≤ 5 × 10^−8^/6 for cross-sex meta-analyses and pooled-sample GWASs. Sex-specific GWAS plots used P ≤ 5 × 10^−8^/12, accounting for six traits in each sex.

### Genomic locus definition and cross-trait comparisons

Genomic loci were defined using FUMA SNP2GENE (v1.8.2)^57^ with GRCh37 coordinates and the 1000 Genomes Phase 3 European reference panel^58^. The executed FUMA configuration used a significance threshold of 8.3 × 10^−9^, linkage disequilibrium (LD) thresholds of r^2^ = 0.6 and 0.1 for independent significant and lead single-nucleotide polymorphisms (SNPs), respectively, and a 250-kb within-trait locus-merging distance. Reference-panel variants were included with a reference MAF threshold of zero; uploaded variants remained subject to MAF ≥ 0.01. Loci overlapping the extended major histocompatibility complex (MHC; chromosome 6:28,477,797-33,448,354) were excluded from reported counts.

Unique loci across the six traits were obtained by merging same-chromosome FUMA intervals overlapping by at least one base pair, including transitive overlaps, without additional flanking distance. Each merged locus was represented by the constituent FUMA top lead SNP with the smallest P value; ties were resolved by a fixed trait order, position and rsID. Cytogenetic bands were assigned using the UCSC hg19 cytoBand annotation. Within each biomarker source, locus sharing was expressed as the proportion of BAU loci overlapping a BAG locus. Regional comparisons used each trait’s smallest available P value within the merged interval, retaining missing results as missing. Shared significant variants were matched by position and unordered allele pair, and selected effect-direction comparisons were aligned to the same effect allele.

### SNP heritability and statistical inflation

Observed-scale SNP heritability was estimated using univariate LD score regression (LDSC v1.0.1) for the sex-specific, cross-sex meta-analysis and pooled-sample GWASs^59^. Summary statistics were harmonized to HapMap3, excluding non-SNP, strand-ambiguous, duplicate and allele-mismatched records. Munging used signed effect directions and P values, with INFO ≥ 0.8 and MAF ≥ 0.01 where measured fields were available. P values stored as zero were replaced by 10^−300^. Analyses used the European reference LD scores and regression weights distributed with LDSC, with freely estimated intercepts. Heritability precision was summarized as:

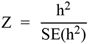

The 95% confidence intervals (CIs) were calculated as:

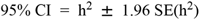

Statistical inflation was described by the LDSC intercept, mean chi-squared statistic and attenuation ratio, calculated as:

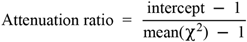

Genomic inflation factors (λ_GC_) were calculated from the one-degree-of-freedom association chi-squared statistics as:

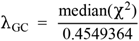

Values calculated from the full QC-passing variant set were distinguished from those obtained from the LDSC SNP subset.

Genetic correlations involving traits with heritability Z < 4, including proteomics-derived BAU, were treated as exploratory.

### Genetic correlations among BAG and BAU traits

Correlations among the six traits were estimated using multivariate LDSC in GenomicSEM (v0.0.5) under R (v4.4.3)^60^, yielding the genetic covariance matrix S and its sampling covariance matrix V. Inputs were aligned to 1,173,036 shared, non-palindromic HapMap3 SNPs with finite Z statistics and positive sample sizes. Chromosome 6:25-34 Mb was excluded. Before SNP intersection, each input was filtered to remove Z^2^ > max (80, 0.001 × maximum per-variant N). Analyses used European LD scores, freely estimated intercepts and 200 genomic jackknife blocks. Genetic correlations (*r_g_*) were calculated by dividing genetic covariance by the square root of the product of the two genetic variances. SEs were obtained by the delta method using the full V matrix. Two-sided tests of *r_g_* = 0 were corrected across the 15 unique pairs using the Benjamini-Hochberg (BH) false discovery rate (FDR) procedure.

### Genetic correlations with external phenotypes

The external library contained 2,936 association datasets: 2,754 FinnGen R13 phenotypes, 78 Psychiatric Genomics Consortium (PGC) datasets from 18 releases, four epigenetic-ageing datasets, eight datasets from the Foote frailty study, three CKDGen datasets, three COVID-19 Host Genetics Initiative (HGI) datasets, one GIANT dataset and 85 additional ageing and health datasets. These were association datasets rather than independent cohorts. Dataset-level publications, accessions, releases, ancestry, sample-size definitions, and download links are recorded in Supplementary Tables 37-38 and Supplementary Note 7.

External statistics were standardized to SNP identifier, alleles, signed effect, SE, P value and sample size. Odds ratios were log-transformed, and signed effect fields were derived from Z statistics when necessary. LDSC munging used P values and signed effect directions and HapMap3 allele alignment. Palindromic variants were excluded.

Bivariate LDSC estimated correlations between each BAG/BAU trait and each external phenotype using European LD scores and regression weights^61^. Intercepts were freely estimated to accommodate correlated errors, including those arising from sample overlap. Numerical eligibility required finite *r_g_* within [−1, 1], positive finite SE, valid P values and positive external heritability where available. A dataset failing these checks for any target was excluded from the global testing family, leaving 2,276 datasets and 13,656 comparisons. Two-sided tests of *r_g_* = 0 underwent global BH correction across this family. Exploratory flags identified low heritability precision (Z < 4), *r_g_* SE > 0.2, fewer than 200,000 shared SNPs, incomplete autosomal coverage, and reconstructed summary statistics.

### Conditional genetic associations of BAG and BAU

To assess associations after mutual adjustment, we regressed each external phenotype’s genetic component jointly on BAG and BAU derived from the same biomarker source. Models covered 45 selected phenotypes for each of the three sources. Three-trait genetic covariance matrices were estimated using the shared-SNP filters, MHC exclusion, free intercepts and 200-block jackknife described for the six-trait analysis. Variables were standardized by their total genetic variances. The standardized genetic regression coefficients for BAG and BAU, mutually adjusted for each other, were:

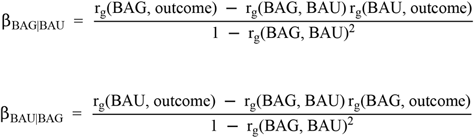

The first equation gives the coefficient for BAG adjusted for BAU; the second gives the coefficient for BAU adjusted for BAG.

Coefficient sampling covariance was JVJ^T^, where J is the analytic Jacobian with respect to the three genetic variances and three genetic covariances. SEs were obtained from the coefficient sampling variances, and 95% CIs used estimate ± 1.96 SE. Two-sided Wald tests were BH-corrected separately across 135 BAG and 135 BAU coefficients. A pooled correction across all 270 coefficients was used as a sensitivity analysis. Estimable models containing any phenotype with heritability Z < 4 were exploratory, including all estimable models containing proteomics-derived BAU.

### Tissue and cell-type enrichment

MAGMA (v1.08) tested gene associations using the SNP-wise mean model, per-variant sample sizes and the 1000 Genomes Phase 3 European LD reference^31^. SNPs were mapped to NCBI37.3 protein-coding gene bodies without flanking extensions, excluding genes overlapping the extended MHC. Significance was assessed using Bonferroni correction across tested genes within each trait. MAGMA tissue-expression analyses used GTEx v8 data for 54 tissues provided through FUMA^62^. One-sided gene-property tests assessed whether higher tissue expression predicted stronger gene associations, accounting for gene–gene correlation. Models adjusted for mean expression across tissues, gene size, gene density and inverse mean minor-allele count, including log-transformed terms for the latter three. BH correction covered all 324 tests across six traits and 54 tissues.

DEPICT (v1-rel194) evaluated 209 tissue, cell-population and biospecimen categories using GPL570 expression data and MeSH annotations^32^. QC-passing variants with P ≤ 5 × 10^−8^/6 were clumped using PLINK^63^ (r^2^ = 0.1; 500 kb). LD estimates used the 1000 Genomes Phase 1 CEU, GBR and TSI reference. Loci were defined using DEPICT’s precomputed LD intervals at an r^2^ threshold of 0.5, extended to gene boundaries and merged. The MHC region (chromosome 6:25-35 Mb) was excluded. Enrichment used 500 gene-count-matched permutations and 50 null repetitions to estimate within-trait FDR, with significance defined as DEPICT-reported FDR < 0.05. Primary analyses required at least ten loci and included the four RBT- and NMR-derived traits. Sensitivity analyses used P ≤ 5 × 10^−8^; traits with fewer than ten primary loci were additionally examined at P ≤ 10^−5^ as exploratory analyses.

### Spatial genetic mapping

We applied gsMap (v1.73.8)^64^ to the Mouse Organogenesis Spatiotemporal Transcriptomic Atlas (MOSTA; STDS0000058)^33^. Analyses included all six GWASs and 53 sections from 14 embryos spanning E9.5-E16.5. Sections were processed independently, excluding undetected genes, unannotated spatial bins and anatomical groups containing fewer than 30 bins.

Following normalization and dimensionality reduction, gsMap integrated gene expression, spatial coordinates and anatomical labels using a graph-attention autoencoder. Gene specificity scores measured local expression enrichment relative to the section-wide background, using neighbouring bins with similar expression profiles and matching anatomical labels. Mouse genes were mapped to human homologues, excluding unmapped and mitochondrial genes.

Gene specificity scores were linked to SNPs using gsMap’s precomputed SNP-gene mappings. Spatial enrichment was assessed using weighted stratified LDSC with the 1000 Genomes Phase 3 European reference, GENCODE release 46 gene annotations mapped to GRCh37 (v46lift37) and baseline annotations. Variants with nonfinite Z statistics or Z^2^ ≥ max(80, 0.001 × N_max_) were excluded, where N_max_ was the maximum per-variant sample size. One-sided tests used HLA-excluded HapMap3 regression weights, freely estimated intercepts and 200 jackknife blocks. Bin-level P values were combined within each anatomical label and section using gsMap’s equal-weight Cauchy procedure with default outlier filtering. Global BH correction was applied across all 5,850 section-tissue-trait tests.

To assess the reproducibility of gsMap associations, we compared tissue-association profiles between the two E16.5 embryos. For each trait and tissue, association scores were calculated as the median section-level -log_10_(Cauchy P) within each embryo. Pearson and Spearman correlations were calculated across 28 shared anatomical labels.

### Single-cell genetic mapping

Single-cell analyses used Tabula Sapiens v2 with author-provided cell annotations and donor metadata^65^. We used the decontXcounts layer, containing decontaminated 10x counts and unchanged Smart-seq counts. Cells required at least 200 detected genes, and genes required detection in at least 50 cells. ERCC spike-ins and ambiguous gene symbols were excluded. Counts were normalized to 10,000 endogenous counts per cell and log1p-transformed. The retained atlas comprised 1,136,218 cells from 24 donors and 28 tissues.

We used scDRS (v1.0.3) to assess enrichment of trait-associated gene expression in individual cells relative to matched control gene sets^34^. For each trait, the top 1,000 genes from complete MAGMA rankings were selected after mapping Entrez identifiers to unambiguous human gene symbols. Gene sets were then intersected with the expression matrix without replacing absent genes. MAGMA P values were converted to association Z-score weights using scDRS’s gene-set preparation procedure. Scoring used scDRS’s default variance-stabilizing weights and 1,000 control gene sets matched on mean expression and variance. Analyses adjusted for sequencing assay and log-transformed detected-gene count. All six traits used the same full-atlas expression background. BH correction was applied across individual-cell P values separately for each trait.

Group analyses included tissue, tissue-by-original-cell-type, broad-compartment and unified-cell-type categories. A fixed crosswalk harmonized 182 original labels into 80 unified cell types, pooling cells across tissues (Supplementary Table 39). Groups required at least 50 cells. For each group, the 95th percentile of normalized scDRS scores was compared with corresponding statistics from the same cells across 1,000 matched control gene sets. Empirical P values and association Z scores were derived from this control distribution. Group-level P values were BH-corrected across all six traits separately for tissue (168 tests), tissue-by-original-cell-type (3,042), unified-cell-type (450) and broad-compartment (36) analyses. Comparisons of BAG and BAU group scores were descriptive, without formal between-trait testing.

### Gene prioritization

We integrated positional, eQTL and chromatin-interaction mapping, MAGMA gene associations and multi-SNP summary-data-based Mendelian randomization (SMR-multi). FUMA positional mapping used a 10-kb window, whereas eQTL mapping applied resource-specific significance thresholds. Chromatin interactions required FDR ≤ 10^−6^, with promoters defined from 250 bp upstream to 500 bp downstream of transcription start sites. Enhancer and promoter filters used Roadmap epigenomic annotations^66^. MAGMA support required Bonferroni significance across tested genes. SMR-multi integrated GWAS associations with expression, splicing and methylation QTLs^67,68^. QTL resources comprised GTEx v8, BrainMeta v2, CAGE, Geuvadis EUR373, PsychENCODE HCP100, Westra, Brain-mMeta and McRae datasets. Summary statistics were obtained from the SMR data resource (https://yanglab.westlake.edu.cn/software/smr/). Analyses used the 1000 Genomes Phase 3 European LD reference, an instrument QTL threshold of P ≤ 5 × 10^−8^ and MAF ≥ 0.01. Support required SMR-multi q < 0.05 after global BH correction across all six traits and resources, together with HEIDI P > 0.01.

The extended MHC was excluded by gene overlap for FUMA and MAGMA and top-variant position for SMR. Each method contributed at most one support indicator per gene, and genes supported by at least three methods were prioritized. BAG-BAU overlap was assessed using standardized gene symbols.

### Functional enrichment analysis

Prioritized genes underwent one-sided hypergeometric over-representation tests using Enrichr collections for GO Biological Process, KEGG and DisGeNET^69,70^. The primary background was the 20,277-gene union of the three collections, restricted separately to genes annotated in each database. Candidate lists and gene sets were intersected with the corresponding database-specific background before enrichment testing. Fold enrichment compared candidate-gene and background proportions within each gene set. Global BH correction covered all tests across six traits and three databases, including zero-overlap terms; significance was q ≤ 0.05.

Sensitivity analyses used the 17,968 genes tested by MAGMA across all six traits as the background, required MAGMA or SMR-multi support within the three-method criterion, or combined both restrictions. Each analysis received separate, complete-family BH correction. In Fig. 5c, the heatmap displayed 30 terms supported in at least one displayed trait across the primary and all three sensitivity analyses.

### Statistical analysis

Statistical tests were two-sided unless otherwise specified. In the molecular and physiological analyses, associations identified in the discovery set were evaluated in the validation set. We used one-sided tests to assess whether these associations were replicated in the same direction. Genetic enrichment tests were one-sided where a positive-enrichment alternative was specified. All confidence intervals were 95%. Heritability estimates, genetic correlations and standardized genetic regression coefficients were reported with standard errors, and their confidence intervals were calculated as the estimate ±1.96 standard errors.

The Benjamini–Hochberg procedure was used for false discovery rate (FDR) correction. In genetic analyses using this correction, significance was defined as q < 0.05, except for over-representation analyses, which used q ≤ 0.05. DEPICT results were evaluated using its native FDR categories. Multiple-testing correction was applied before genetic results were selected for display.

Probabilistic age models were implemented in Python 3.10.19 using NGBoost 0.5.10, LightGBMLSS 0.6.1 and XGBoostLSS 0.5.0, with LightGBM 4.6.0, XGBoost 3.2.0 and scikit-learn 1.7.2. Statistical analyses and data visualization were performed in R 4.6.0.

## Author Contributions

Conceptualization, Y.L., G.F., S.Y., Q.H., J.J., and Q.D.; methodology, Y.L., G.F., S.Y., Q.H., and J.J.; software, Y.L., G.F.; formal analysis, Y.L., G.F., and L.K., with additional analyses performed by S.Y., Q.H., and J.J.; data curation, Y.L., G.F., S.Y., Q.H., J.J., Y.W., Zeyuan Pei, L.X.T., K.L., Zean Pan, and Z.Y.; validation, Y.L., G.F., S.Y., Q.H., J.J., K.G., and Q.D.; visualization, Y.L. and G.F.; resources, Y.W., L.G., and J.S.J.; supervision, L.K., K.G., and Q.D.; project administration, L.K., K.G., and Q.D.; writing - original draft, Y.L. and G.F.; writing - review & editing, all authors.

The corresponding author attests that all listed authors meet authorship criteria and that no others meeting the criteria have been omitted. This work was a collaborative team effort. The order of co-first authors reflects the authors’ agreed-upon arrangement. All co-first authors contributed equally to this work. When citing this article for professional or academic purposes, each co-first author may present the co-first author group with their own name listed first.

## Acknowledgement

We acknowledge the research support from the National Key Research and Development Program of China (number 2024YFC3607002); Key Research and Development Project of the Science and Technology Department of Xinjiang Uygur Autonomous Region, No. 20253149440, (Project Number: 20253150247); Key Research and Development Program Projects of Xinjiang Uygur Autonomous Region, No. 2025B3016, (Project Number: 2025B3016-3); National Natural Science Foundation of China (number 42277419); Capital’ s Funds for Health Improvement and Research (No 2026-1Q-1012); and the Research Fund of Vanke School of Public Health in Tsinghua University. The funders had no role in study design, data collection and analysis, decision to publish or preparation of the manuscript.

## Data availability

The UK Biobank data analyzed in this study are available to eligible researchers through the UK Biobank Access Management System following approval of a research application (https://www.ukbiobank.ac.uk/use-our-data/apply-for-access/). This study was conducted under UK Biobank approved applications 75587 and 65036, with most analyses completed under application 75587 and subsequent analyses conducted under application 65036. The NHANES 1999-2018 survey data and the corresponding 2019 public-use linked mortality files are available from the US National Center for Health Statistics (https://wwwn.cdc.gov/nchs/nhanes/continuousnhanes/;https://www.cdc.gov/nchs/linked-data/mortality-files/). The CHARLS 2011 baseline and follow-up data are available to registered researchers through the CHARLS website following acceptance of the data-use agreement and approval of a data request (https://charls.pku.edu.cn/en). The CHNS 2009 biomarker and follow-up data are available as household- and individual-level datasets through the UNC Dataverse (https://chns.cpc.unc.edu/data/datasets/). Hallmark, Reactome and Gene Ontology gene sets from MSigDB v2026.1.Hs are available after free registration (https://www.gsea-msigdb.org/gsea/msigdb/index.jsp).

## Code availability

The codes used for data preprocessing, model training, calibration and inference, validation, statistical analyses and figure plots is available on GitHub at https://github.com/AI4HEALTH-LAB-THU/BAU. Software dependencies and instructions for reproducing the analyses are provided in the repository.

## Supplementary information

Supplementary Information includes: **Supplementary Figures 1-9, Supplementary Tables 1-39, Supplementary Notes 1-7.**

---

**Extended Data Fig. 1.**
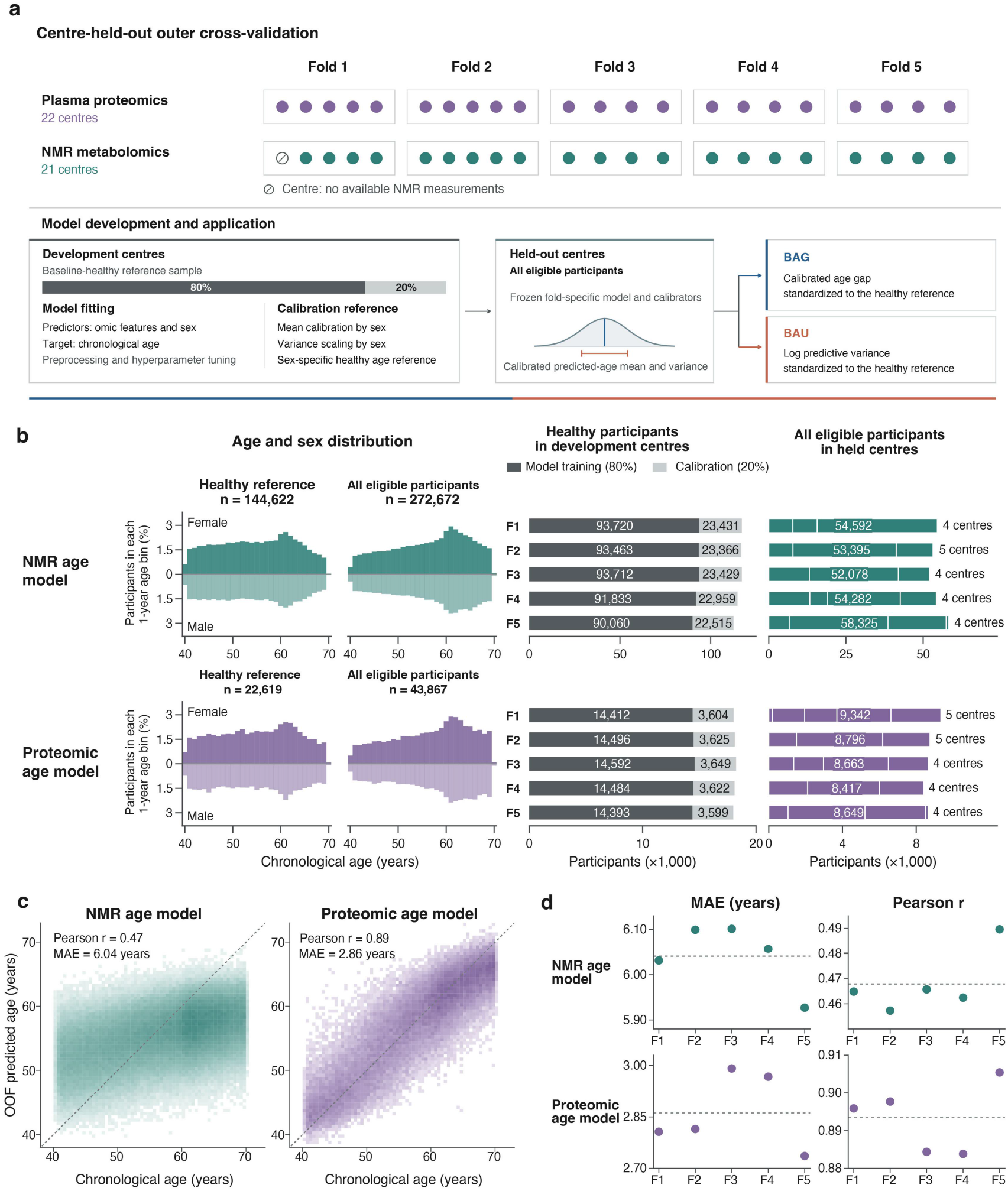
Development and validation of probabilistic biological age models. **a,** Model development and validation across five folds defined by UK Biobank assessment centres. Each dot represents one centre, with 22 centres contributing proteomic data and 21 contributing NMR data. For each fold, healthy participants from the remaining centres were divided into model-fitting (80%) and independent calibration (20%) samples. Models used molecular measurements and sex to predict chronological age. After calibration, models were applied to all eligible participants in the held-out centres. BAG and BAU were derived from the calibrated age gap and log predictive variance, respectively, and standardized relative to age- and sex-specific healthy references. **b,** Sample composition for the NMR and proteomic models. Left, age and sex distributions of the healthy-reference and all-eligible populations. Bar heights show the percentage of the total population represented by each combination of sex and 1-year age interval, with female participants shown in the upper half and male participants in the lower half. Middle, model-fitting and calibration sample sizes in each fold. Right, sample sizes in held-out centres. **c,** Age prediction performance in held-out participants. Predicted age is the calibrated mean of each participant’s predicted-age distribution, obtained while their assessment centre was held out of model development. Darker colours indicate more participants. Pearson correlations and mean absolute errors (MAEs) are shown. OOF, out-of-fold. **d,** Age prediction performance across the five held-out folds (F1-F5). Points show fold-specific MAEs and Pearson correlations. Horizontal dashed lines show the corresponding estimates calculated from all out-of-fold predictions combined. NMR, nuclear magnetic resonance.

**Extended Data Fig. 2.**
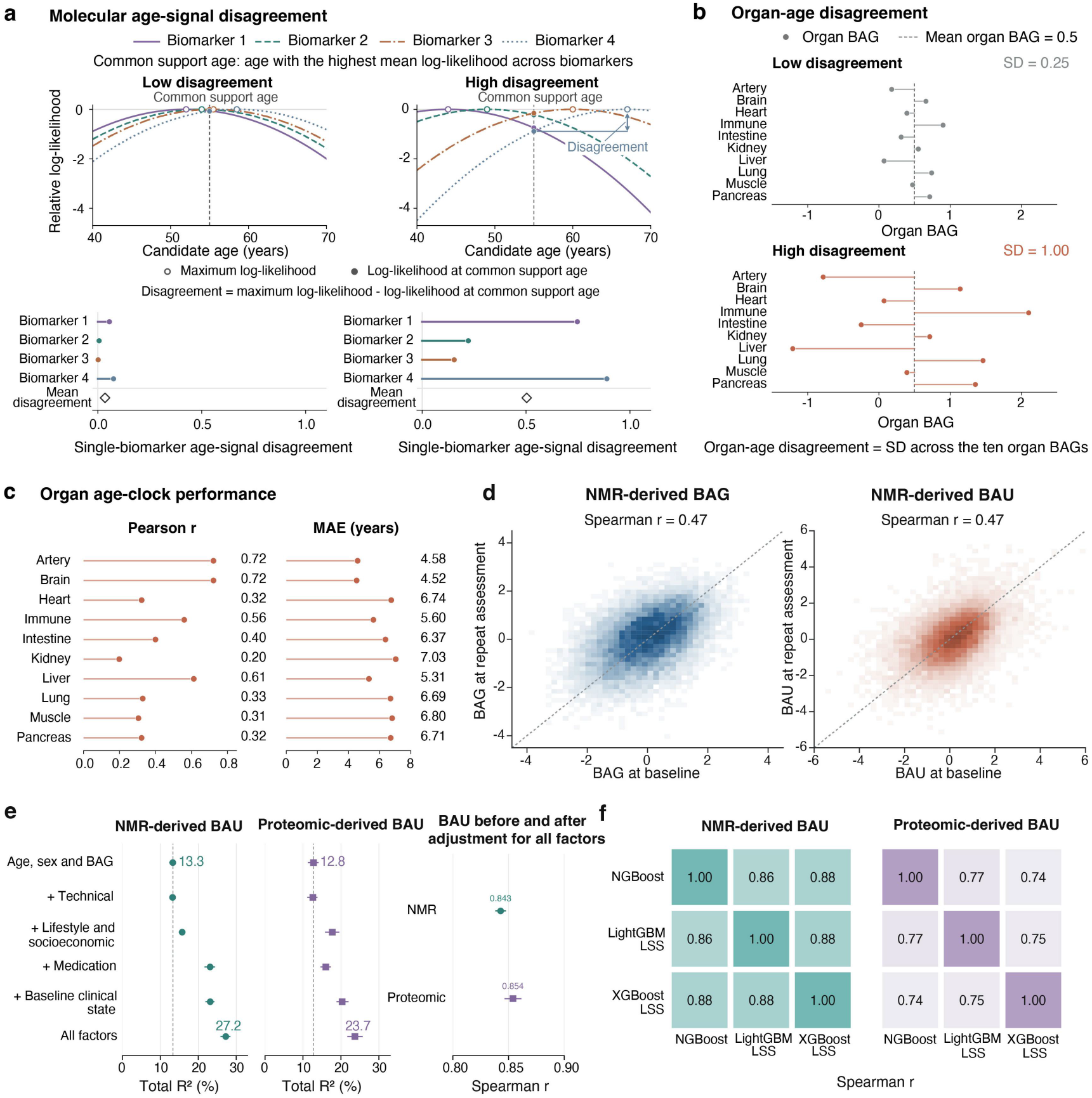
Molecular and organ-age disagreement and robustness of BAU. **a,** Illustration of molecular age-signal disagreement using four biomarkers. Likelihood measures how well a biomarker’s observed level matches the levels expected at a given age in the healthy reference population. Curves show log-likelihood relative to each biomarker’s maximum. The common support age maximizes the mean log-likelihood across biomarkers. Open circles mark individual maxima, and filled circles mark values at the common support age. Their difference defines single-biomarker disagreement. Lower plots show these differences and their mean (diamonds) for examples with low and high disagreement. This calculation was applied separately to proteins and NMR measures. **b,** Illustration of organ-age disagreement using ten standardized organ BAGs. The two examples have the same mean organ BAG but different variation across organs. Points represent individual organ BAGs, and horizontal segments connect them to the mean (dashed line). Organ-age disagreement is the standard deviation across the ten organ BAGs. **c,** Prediction performance of the ten proteomic organ clocks used to calculate organ-age disagreement. Points show Pearson correlations between predicted and chronological age (left) and mean absolute errors (MAEs; right), calculated from all available held-out predictions for each organ. **d,** Longitudinal persistence of NMR-derived BAG and BAU in 13,620 participants with a median interval of 4.36 years between baseline and reassessment. BAG and BAU at reassessment were calculated using the same models and calibration parameters used at baseline. Spearman correlations between baseline and reassessment values are shown for BAG and BAU separately. **e,** Variation in BAU explained by measured factors and preservation of participant rankings after adjustment. Left and middle, total R² for predicting NMR-derived and proteomic-derived BAU in held-out assessment centres. The reference model included age, sex and BAG. Each additional factor group was added separately to this model; the all-factor model included technical, lifestyle and socioeconomic, medication and baseline clinical factors together. Vertical dashed lines mark reference-model R². Right, Spearman correlations between original and adjusted BAU. For each participant, BAU was predicted from age, sex, BAG and all four factor groups using a model trained in other assessment centres. Adjusted BAU was calculated as the participant’s original BAU minus this predicted value. Points and horizontal lines show estimates and 95% confidence intervals. **f,** Correlations between BAU estimates from NGBoost, LightGBMLSS and XGBoostLSS. All three algorithms used the same assessment-centre folds and healthy-reference calibration procedure. Cells show pairwise Spearman correlations between their BAU estimates in the same participants, separately for NMR-derived BAU (n = 272,672) and proteomic-derived BAU (n = 43,867). Darker colours indicate higher correlations. NMR, nuclear magnetic resonance.

**Extended Data Fig. 3.**
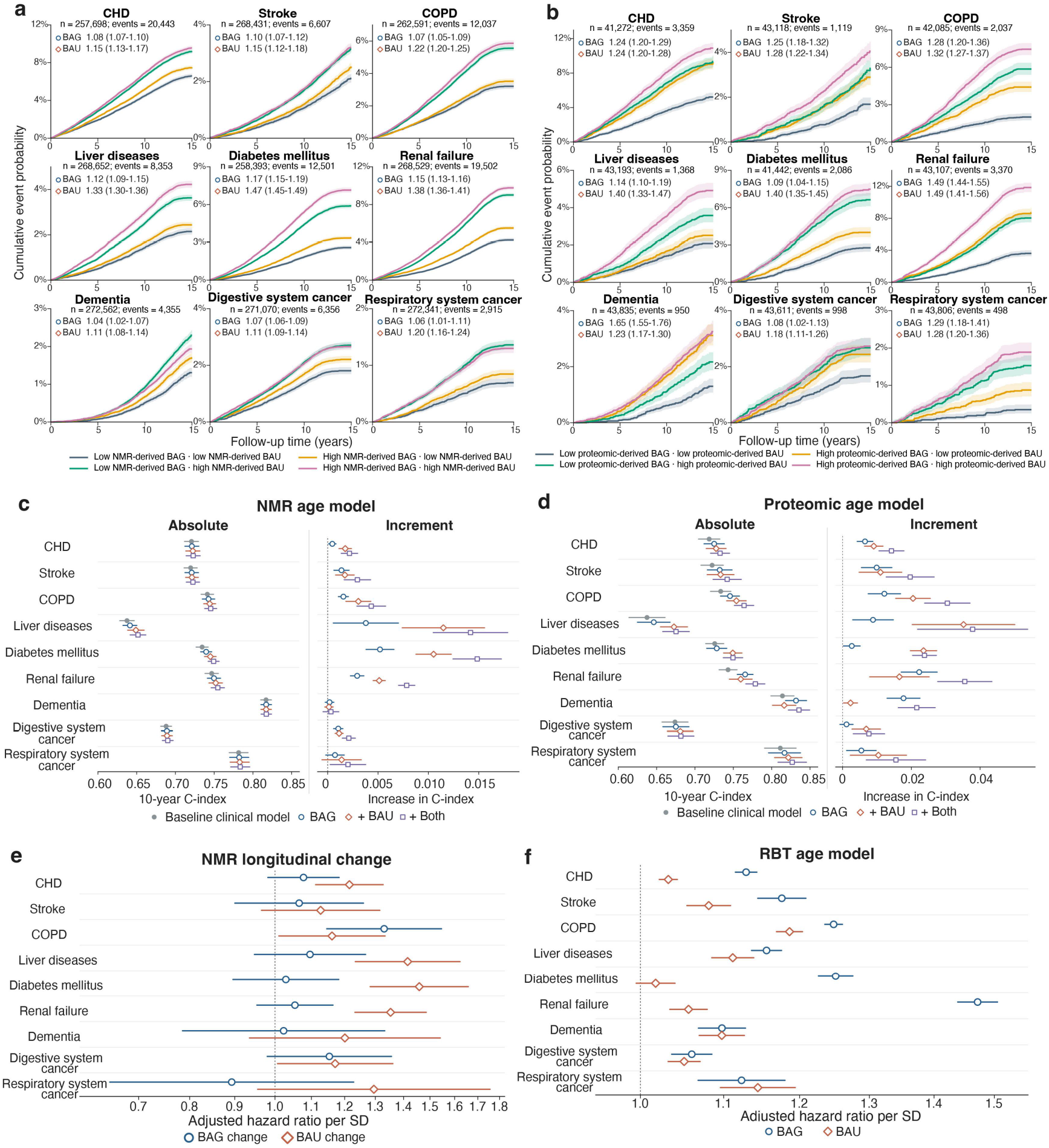
Disease-specific associations and risk prediction using BAU and BAG. **a,** Fifteen-year cumulative probabilities of nine incident diseases in four groups defined by low or high NMR-derived BAG and BAU. Low and high values were defined as <0 and ≥0 on the healthy-reference scale, respectively. Each disease analysis included participants free of that disease at baseline. Curves show one minus Kaplan-Meier estimates, and shading indicates 95% confidence intervals. Values within each plot are adjusted hazard ratios (HRs) and 95% confidence intervals per 1 SD higher BAG or BAU. Cox models included BAG and BAU together and adjusted for age, sex, smoking, alcohol intake, physical activity, education, income and employment, with baseline hazards stratified by assessment centre. Death before disease onset was treated as censoring. **b,** As in **(a)**, for proteomic-derived BAG and BAU. **c,** Ten-year C-index and its increase after adding NMR-derived BAG, BAU or both to a baseline clinical model for each disease. The baseline model included age, sex, smoking, diabetes history, systolic blood pressure, total cholesterol, HDL cholesterol and antihypertensive treatment. Left, C-index for the baseline and expanded models. Right, the difference in C-index between each expanded model and the baseline model. Predictions were evaluated in five folds with separate assessment centres used for training and validation. Points and horizontal lines show estimates and 95% confidence intervals. **d,** As in **(c)**, for proteomic-derived BAG and BAU. **e,** Associations of changes in NMR-derived BAG and BAU with the nine incident diseases. Change was calculated as the reassessment value minus the baseline value. Follow-up began at reassessment, with each analysis including participants free of the corresponding disease at that visit. Cox models included baseline BAG and BAU and both change measures, with adjustment for age at reassessment, the interval between assessments and the remaining covariates in **(a)**. Death before disease onset was treated as censoring. Points and horizontal lines show HRs per 1 SD of change and 95% confidence intervals. **f,** Associations of RBT-derived BAG and BAU with the nine incident diseases in UK Biobank. Cox models were specified as in **(a)**. Points and horizontal lines show adjusted HRs per 1 SD and 95% confidence intervals. CHD, coronary heart disease; COPD, chronic obstructive pulmonary disease; HDL, high-density lipoprotein; NMR, nuclear magnetic resonance; RBT, routine blood tests.

**Extended Data Fig. 4.**
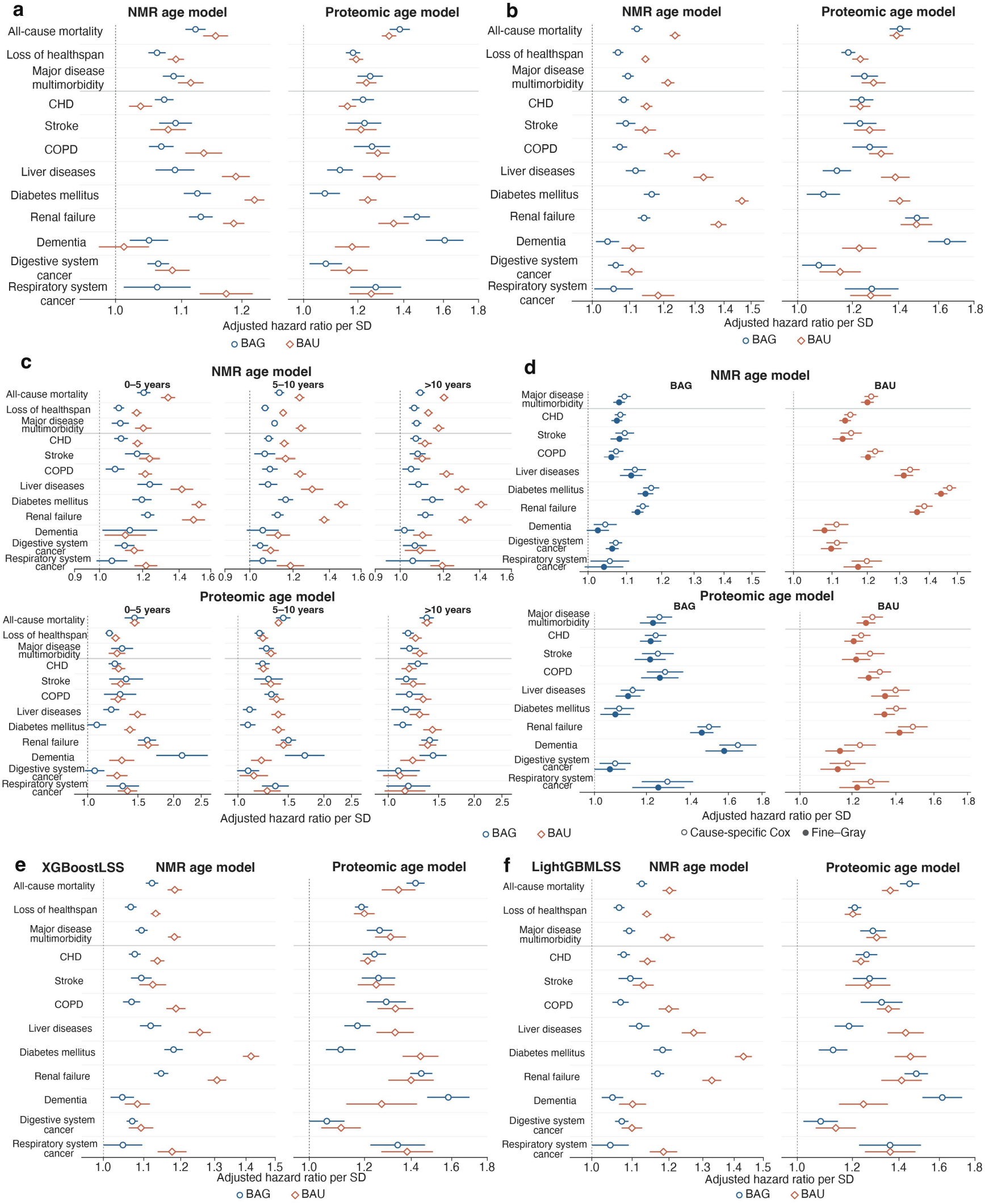
Sensitivity analyses of associations between BAG, BAU and health outcomes. **a,** Associations of NMR-derived and proteomic-derived BAG and BAU with three primary outcomes and nine incident diseases after extended covariate adjustment. Primary Cox models included BAG and BAU together and adjusted for age, sex, smoking, alcohol intake, physical activity, education, income and employment. Extended models additionally included diet, medication use, cardiovascular disease and diabetes family history, body mass index, systolic blood pressure and baseline disease burden. Medications included lipid-lowering drugs, antihypertensive drugs and insulin. Disease burden was classified as 0, 1 or ≥2 of 23 baseline conditions. Points and horizontal lines show adjusted hazard ratios (HRs) per 1 SD and 95% confidence intervals. **b,** Associations after excluding events during the first year following baseline. Participants were required to be alive, under follow-up and free of the target outcome at one year. Follow-up began at that time, and Cox models used the primary adjustment set. **c,** Associations of baseline BAG and BAU with outcomes during 0-5, 5-10 and >10 years of follow-up, shown for NMR-derived measures (top) and proteomic-derived measures (bottom). Separate Cox models were fitted for each interval using the primary adjustment set. Participants entering later intervals were alive, under follow-up and free of the target outcome at the start of that interval. **d,** Associations with major disease multimorbidity and nine incident diseases under alternative approaches to competing mortality. Open circles show cause-specific Cox HRs, with death before the target event treated as censoring. Filled circles show Fine-Gray subdistribution HRs, with death treated as a competing event. Both models included BAG and BAU together and used the primary adjustment set. Horizontal lines indicate 95% confidence intervals. **e,** Associations with the same 12 outcomes using BAG and BAU generated by XGBoostLSS. The primary Cox analyses were repeated using these measures, separately for NMR and proteomic models. Points and horizontal lines show adjusted HRs per 1 SD and 95% confidence intervals. **f,** As in **(e)**, using BAG and BAU generated by LightGBMLSS. CHD, coronary heart disease; COPD, chronic obstructive pulmonary disease; NMR, nuclear magnetic resonance.

**Extended Data Fig. 5.**
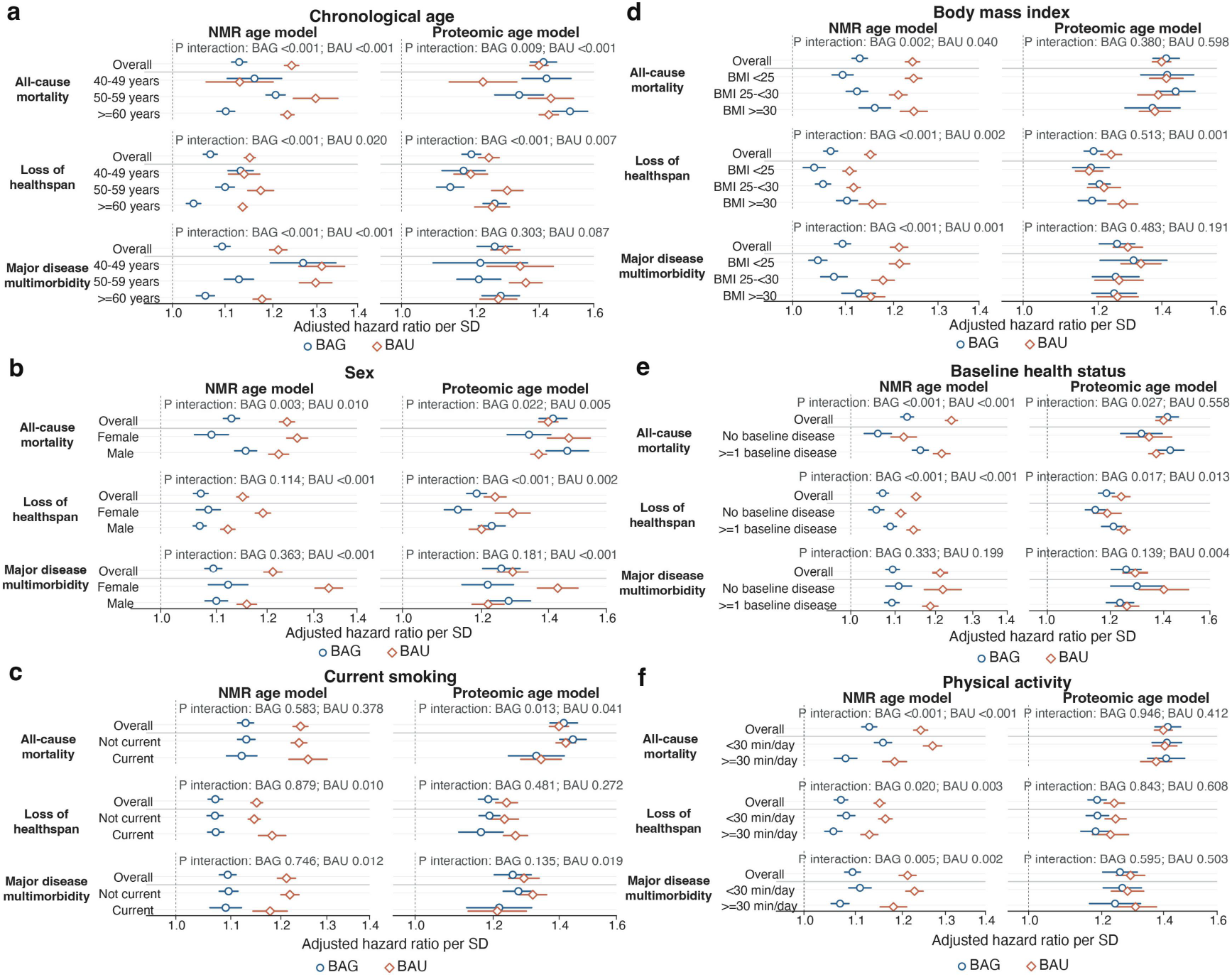
Associations of BAG and BAU across participant subgroups. **a-f,** Subgroup analyses assessing whether associations of BAG and BAU with all-cause mortality, loss of healthspan and major disease multimorbidity differ by age **(a)**, sex **(b)**, current smoking **(c)**, body mass index **(d)**, baseline health status **(e)** and physical activity **(f)**. Results are shown separately for NMR-derived and proteomic-derived BAG and BAU. Age groups were 40-49, 50-59 and ≥60 years; BMI groups were <25, 25-<30 and ≥30 kg m⁻². Baseline health status distinguished participants with none versus at least one of 23 conditions. Physical activity groups were defined by daily moderate-to-vigorous activity of <30 or ≥30 minutes. Overall rows show associations in all eligible participants for each outcome, estimated from the primary Cox models. Subgroup HRs were estimated from Cox models including BAG, BAU, the subgroup variable and its interactions with both BAG and BAU. Models adjusted for age, sex, current smoking, alcohol intake, physical activity, education, income and employment, with baseline hazards stratified by assessment centre. Multimorbidity models were additionally stratified by baseline disease-domain count. Circles and diamonds show adjusted HRs for BAG and BAU, respectively, per 1 SD on the healthy-reference scale; horizontal lines indicate 95% confidence intervals. Interaction P values test whether the BAG or BAU association differs across subgroups and were obtained from global Wald tests. The figure shows P values before correction for multiple testing. BMI, body mass index; NMR, nuclear magnetic resonance.

**Extended Data Fig. 6.**
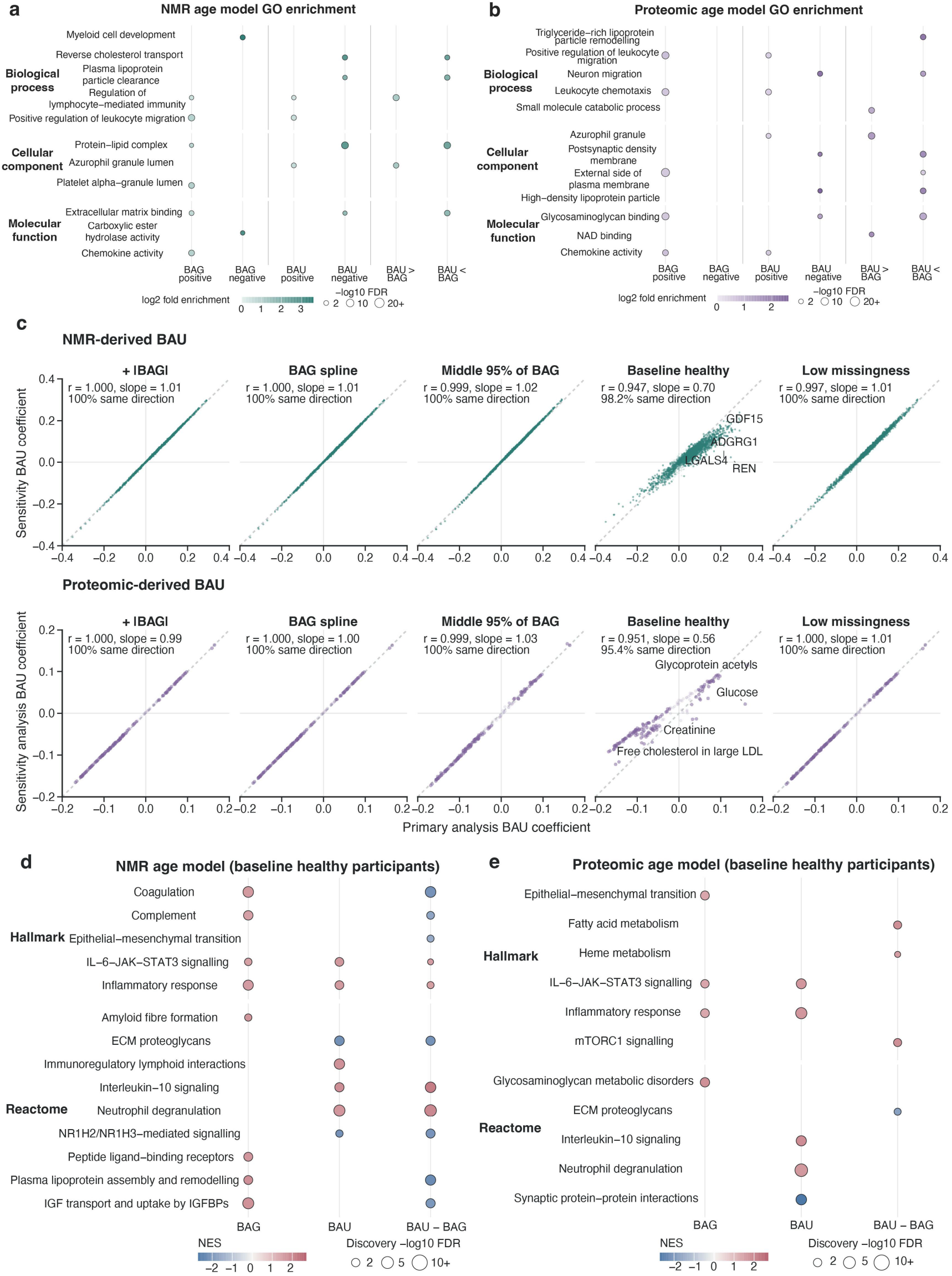
Functional annotation and sensitivity analyses of molecular associations with BAG and BAU. **a,** Gene Ontology (GO) over-representation analysis of proteins with replicated associations with NMR-derived BAG or BAU, or replicated differences between their association coefficients. The six columns represent positive and negative BAG associations, positive and negative BAU associations, and positive and negative coefficient differences (βBAU−βBAG). Enrichment was tested against the measured proteins mapped to each GO category, using one-sided hypergeometric tests. Benjamini-Hochberg correction was applied separately within each protein group and GO category. Representative terms with FDR <0.05 are shown. Colour indicates log2 fold enrichment, and bubble area indicates -log10 FDR. **b,** As in **(a)**, for proteomic-derived BAG and BAU, using 2,715 proteins after excluding the 204 proteins included in the proteomic age model. **c,** Sensitivity of BAU molecular associations to alternative BAG adjustments and participant restrictions. Each point compares a molecule’s association coefficient in the primary analysis (x axis) with its coefficient in a sensitivity analysis (y axis). The upper row shows NMR-derived BAU associations with 2,919 proteins; the lower row shows proteomic-derived BAU associations with 168 NMR measures. From left to right, analyses added absolute BAG to the joint BAG and BAU model, replaced linear BAG with a spline, restricted BAG to its central 95%, restricted participants to those healthy at baseline, or restricted molecular-assay missingness to ≤1%. Pearson correlations and slopes from regressions through the origin were calculated across all molecules. Darker points identify primary associations replicated in held-out centres; percentages indicate the proportion of these associations retaining the same direction. **d,** Hallmark and Reactome enrichment analysis in baseline-healthy participants to assess whether the pathway findings in Fig. 3 persisted in this population. NMR-derived BAG and BAU associations with 2,919 proteins were re-estimated, and proteins were ranked separately by the association statistics for BAG, BAU and their coefficient difference (βBAU−βBAG). Displayed pathways were drawn from those selected for Fig. 3 and had at least one result meeting discovery set FDR <0.05 and same-direction held-out validation FDR <0.05. Colour indicates the discovery set normalized enrichment score (NES), and bubble area indicates discovery set -log10 FDR. **e,** As in **(d)**, for proteomic-derived BAG and BAU associations with 2,715 proteins outside the proteomic age model. FDR, false discovery rate; NMR, nuclear magnetic resonance.

**Extended Data Fig. 7.**
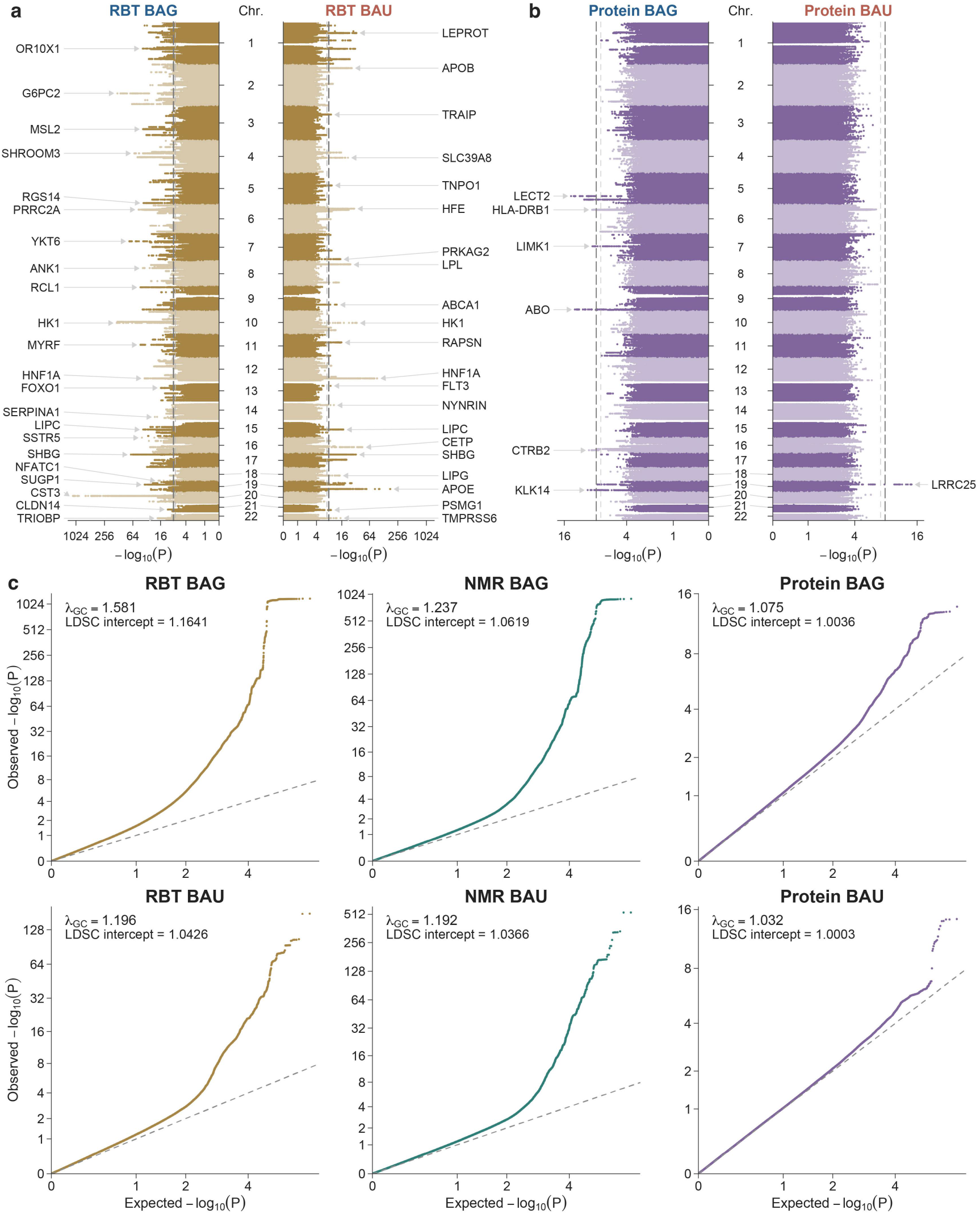
Genome-wide association and quantile-quantile plots for BAG and BAU. **a-b**, Vertical Miami plots of genome-wide association study (GWAS) results for RBT-derived **(a)** and proteomics-derived **(b)** BAG and BAU. BAG and BAU are shown on the left and right, respectively. Horizontal axes display log_2_[1-log_10_(P)], with tick labels showing -log_10_(P). Alternating shades distinguish adjacent chromosomes. Light dashed and dark long-dashed lines indicate P = 5 × 10^−8^ and P = 8.33 × 10^−9^ (5 × 10^−8^/6), respectively. Labels identify the nearest genes to the most significant variant meeting the adjusted threshold on each chromosome. **c**, Quantile–quantile (Q-Q) plots for the six traits derived from RBT, NMR and proteomics. Horizontal and vertical axes show expected and observed -log_10_(P), respectively, using the same transformation and tick-label convention as **(a)** and **(b)**. Each panel reports the genomic inflation factor (λ_GC_) and linkage disequilibrium score regression (LDSC) intercept. All results are from cross-sex fixed-effect inverse-variance GWAS meta-analyses, and association tests were two-sided.

**Extended Data Fig. 8.**
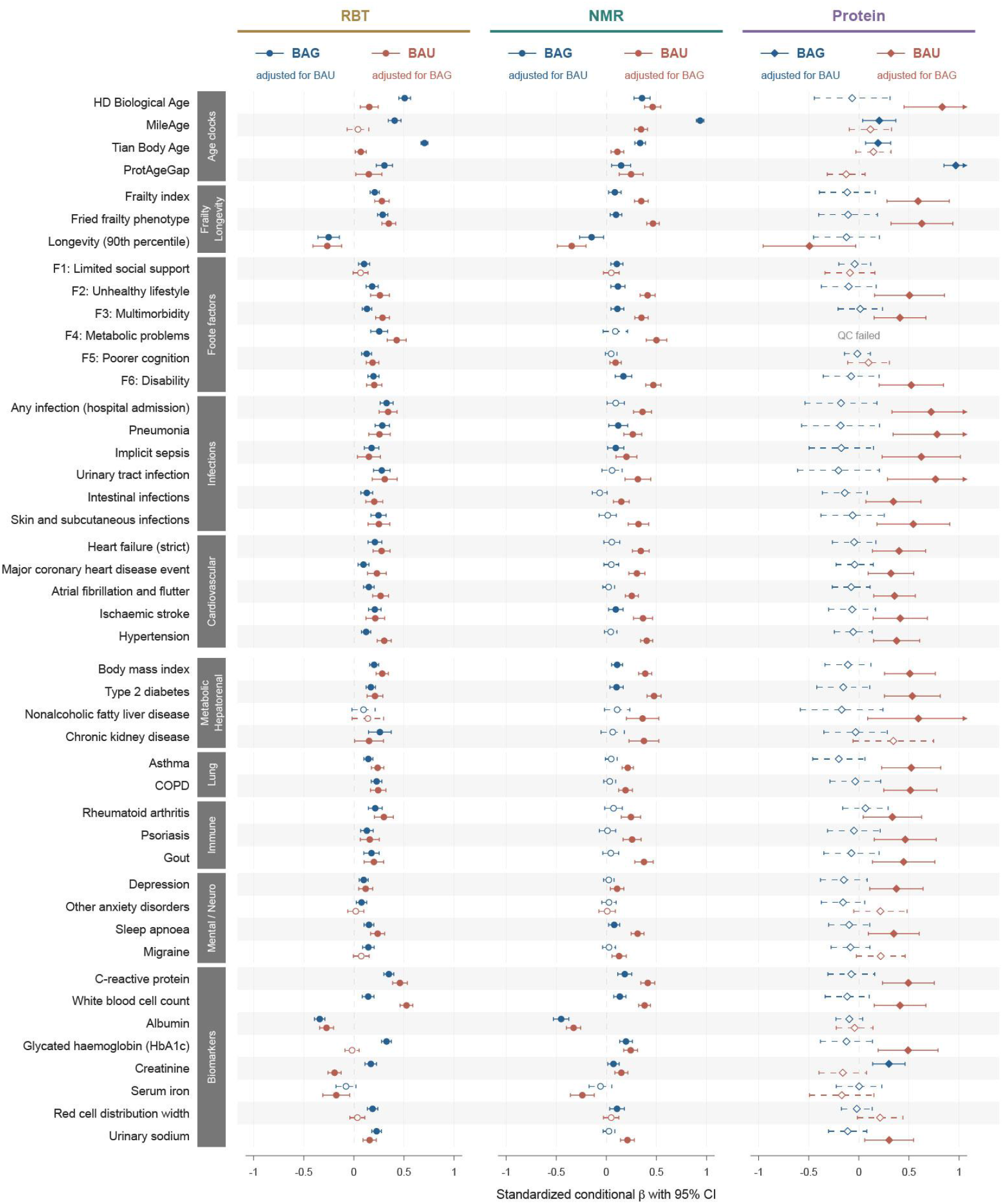
Mutually adjusted genetic associations of BAG and BAU with health-related phenotypes. Standardized conditional genetic regression coefficients (β) with 45 selected external phenotypes, estimated using joint genetic covariance models incorporating BAG and BAU from the same biomarker source. Columns show results for traits derived from RBT, NMR metabolomics and proteomics. Blue denotes BAG adjusted for BAU, and red denotes BAU adjusted for BAG. Points and horizontal bars indicate estimates and 95% confidence intervals (CIs), respectively. Two-sided Wald tests were corrected using the Benjamini-Hochberg (BH) false discovery rate (FDR) procedure, separately for BAG and BAU across 135 tests each. Filled symbols and solid intervals indicate q < 0.05; open symbols and dashed intervals indicate q ≥ 0.05. Diamonds denote exploratory proteomic joint models involving the low-heritability-precision BAU trait. Vertical dashed lines indicate β = 0; arrowheads indicate CIs extending beyond the displayed axis limits. Models failing covariance quality control (QC) are marked as “QC failed”.

**Extended Data Fig. 9.**
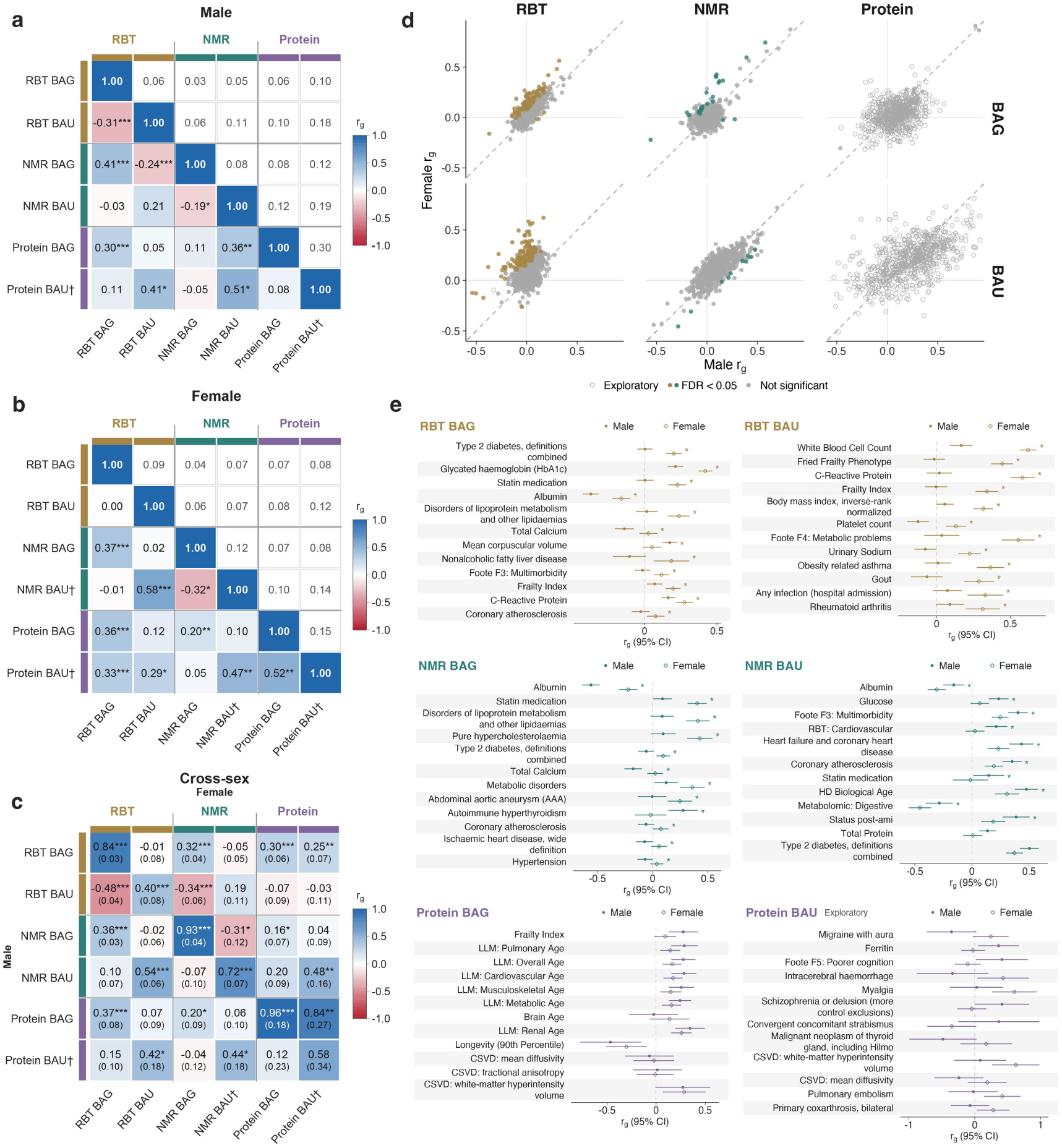
Sex-stratified and cross-sex genetic correlations of BAG and BAU. **a-b**, Genetic correlations (*r_g_*) among the six traits in males **(a)** and females **(b)**, estimated using linkage disequilibrium score regression (LDSC). Lower and upper triangles show r₍g₎ estimates and standard errors (SEs), respectively. **c**, Cross-sex correlations, with male traits in rows and female traits in columns; SEs are shown in parentheses. In a-c, blue and red indicate positive and negative correlations. Asterisks denote Benjamini-Hochberg (BH) false discovery rate (FDR) significance across 66 two-sided tests: q < 0.05 (*), q < 0.01 (**) and q < 0.001 (***). Daggers indicate low heritability precision (Z-score < 4 in univariate or pairwise analyses). **d**, Male versus female genetic correlations with external phenotypes. Coloured and gray filled points indicate significant and nonsignificant sex differences, respectively; open circles denote exploratory estimates. Diagonal dashed lines indicate equal correlations. **e**, Selected sex-specific correlations. Filled circles and open diamonds denote males and females, respectively; horizontal bars indicate 95% confidence intervals. Vertical dashed lines indicate *r_g_* = 0. In **(d-e)**, sex differences were assessed using two-sided Wald tests with paired block-jackknife uncertainty and global BH-FDR correction. Asterisks indicate significant differences between male and female genetic correlations (global BH-FDR q < 0.05). Proteomics-derived BAU comparisons are exploratory.

**Extended Data Fig. 10.**
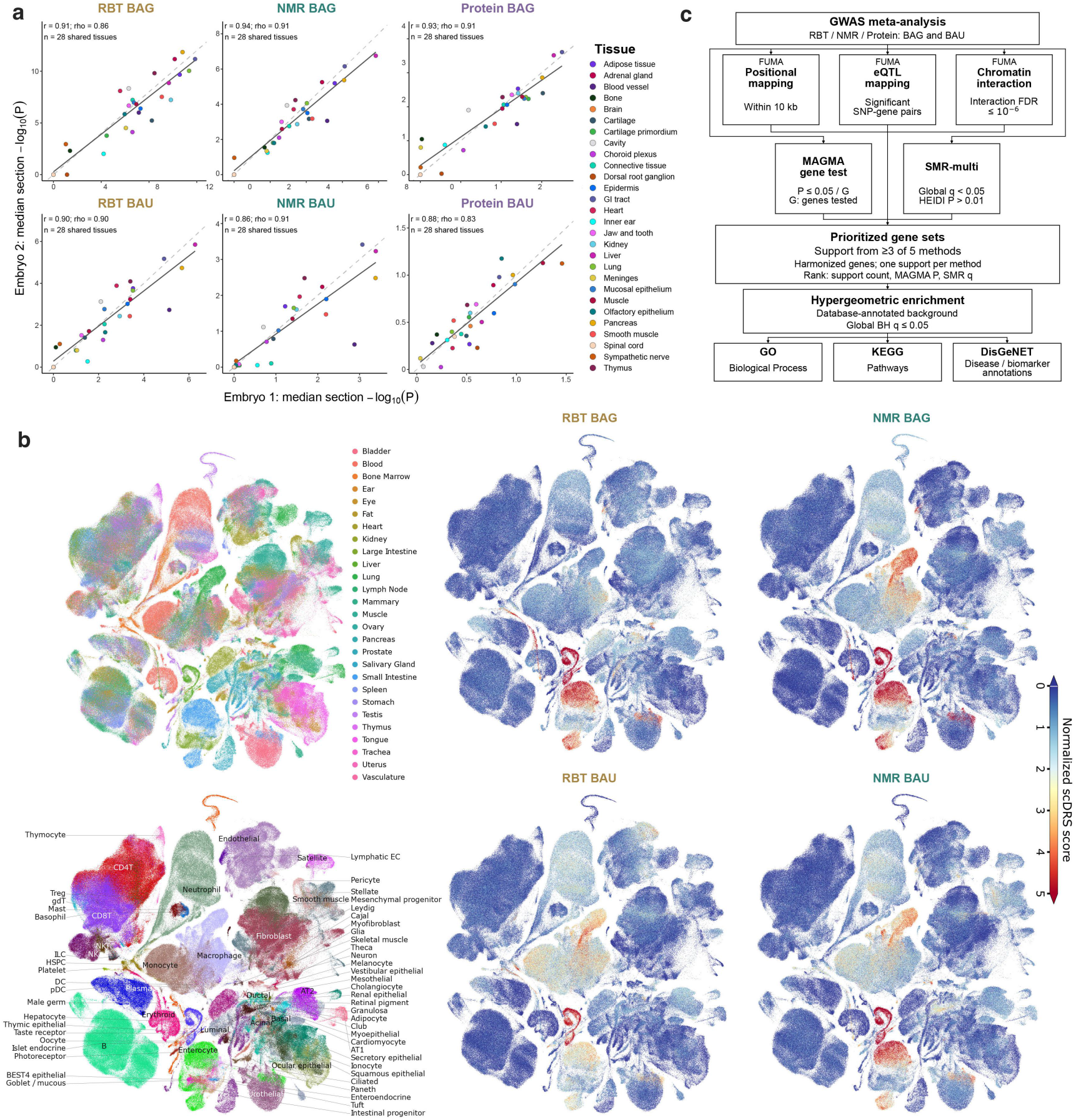
Spatial concordance, single-cell mapping and gene prioritization of BAG and BAU. **a**, Concordance of gsMap tissue-association profiles between two E16.5 mouse embryos. Points represent shared tissues, coloured by tissue identity; axes show median section-level -log_10_(P) values within each embryo. Solid and dashed lines indicate linear fits and equality, respectively. Pearson’s r and Spearman’s ρ summarize profile concordance. **b**, Uniform manifold approximation and projection (UMAP) of the Tabula Sapiens v2 atlas, coloured by tissue, cell type or normalized single-cell disease relevance scores (scDRS) for RBT- and NMR-derived BAG and BAU. Blue-to-red colors indicate increasing scores. **c**, Overview of gene prioritization and downstream functional enrichment analyses.

## References

1. Oh, H.S.-H., et al. Organ aging signatures in the plasma proteome track health and disease. Nature 624, 164–172 (2023).

2. Tian, Y.E., et al. Heterogeneous aging across multiple organ systems and prediction of chronic disease and mortality. Nature Medicine 29, 1221–1231 (2023).

3. Rutledge, J., Oh, H. & Wyss-Coray, T. Measuring biological age using omics data. Nature Reviews Genetics 23, 715–727 (2022).

4. Kroemer, G., et al. From geroscience to precision geromedicine: Understanding and managing aging. Cell 188, 2043–2062 (2025).

5. Horvath, S. & Raj, K. DNA methylation-based biomarkers and the epigenetic clock theory of ageing. Nature Reviews Genetics 19, 371–384 (2018).

6. Belsky, D.W., et al. Quantification of biological aging in young adults. PNAS 112, E4104–4110 (2015).

7. Li, Y., et al. Large language model-based biological age prediction in large-scale populations. Nature Medicine 31, 2977–2990 (2025).

8. Wang, Y., et al. Organ-specific proteomic aging clocks predict disease and longevity across diverse populations. Nature Aging 6, 162–180 (2026).

9. Yang, Z., et al. Brain aging patterns in a large and diverse cohort of 49,482 individuals. Nature Medicine 30, 3015–3026 (2024).

10. Cao, H., et al. MRI-based multi-organ clocks for healthy aging and disease assessment. Nature Medicine 32, 82–92 (2026).

11. Abila, E., et al. Histological aging signatures for monitoring tissue-specific aging and disease. Nature Medicine (2026).

12. Ding, D.Y., et al. Plasma proteomic signatures of cellular aging predict human disease. Nature Medicine 32, 2060–2072 (2026).

13. Tyshkovskiy, A., et al. Universal transcriptomic hallmarks of mammalian ageing and mortality. Nature 654, 173–188 (2026).

14. Kriukov, D., Efimov, E., Gelfand, M.S., Moskalev, A. & Khrameeva, E.E. Do we actually need aging clocks? NPJ Aging (2025).

15. Argentieri, M.A., et al. Proteomic aging clock predicts mortality and risk of common age-related diseases in diverse populations. Nature Medicine 30, 2450–2460 (2024).

16. Moqri, M., et al. Validation of biomarkers of aging. Nature Medicine 30, 360–372 (2024).

17. Wyss-Coray, T. & Topol, E.J. Biological aging clocks in health and disease. Nature Medicine 32, 2383–2394 (2026).

18. Riley, R.D., et al. Uncertainty of risk estimates from clinical prediction models: rationale, challenges, and approaches. BMJ 388(2025).

19. Li, Y., Goodrich, J.M., Peterson, K.E., Song, P.X.K. & Luo, L. Uncertainty quantification in epigenetic clocks via conformalized quantile regression. Genetic Epidemiology 49, e70008 (2025).

20. Bahar, R., et al. Increased cell-to-cell variation in gene expression in ageing mouse heart. Nature 441, 1011–1014 (2006).

21. Martinez-Jimenez, C.P., et al. Aging increases cell-to-cell transcriptional variability upon immune stimulation. Science 355, 1433–1436 (2017).

22. Li, Q., et al. Homeostatic dysregulation proceeds in parallel in multiple physiological systems. Aging Cell 14, 1103–1112 (2015).

23. Scheffer, M., et al. Quantifying resilience of humans and other animals. PNAS 115, 11883–11890 (2018).

24. Richardson, T.G., et al. Effects of apolipoprotein B on lifespan and risks of major diseases including type 2 diabetes: a mendelian randomisation analysis using outcomes in first-degree relatives. Lancet Healthy Longevity 2, e317–e326 (2021).

25. Barzilai, N., et al. Unique lipoprotein phenotype and genotype associated with exceptional longevity. JAMA 290, 2030–2040 (2003).

26. Lin, W., et al. Hepatic metal ion transporter ZIP8 regulates manganese homeostasis and manganese-dependent enzyme activity. The Journal of Clinical Investigation 127, 2407–2417 (2017).

27. Ng, S.W.K., et al. Convergent somatic mutations in metabolism genes in chronic liver disease. Nature 598, 473–478 (2021).

28. Aksentijevich, I., et al. An autoinflammatory disease with deficiency of the interleukin-1-receptor antagonist. The New England Journal of Medicine 360, 2426–2437 (2009).

29. Dewey, F.E., et al. Inactivating Variants in ANGPTL4 and Risk of Coronary Artery Disease. The New England Journal of Medicine 374, 1123–1133 (2016).

30. Cohen, A.A., et al. A novel statistical approach shows evidence for multi-system physiological dysregulation during aging. Mechanisms of Ageing and Development 134, 110–117 (2013).

31. de Leeuw, C.A., Mooij, J.M., Heskes, T. & Posthuma, D. MAGMA: generalized gene-set analysis of GWAS data. PLoS Computational Biology 11, e1004219 (2015).

32. Pers, T.H., et al. Biological interpretation of genome-wide association studies using predicted gene functions. Nature Communications 6, 5890 (2015).

33. Chen, A., et al. Spatiotemporal transcriptomic atlas of mouse organogenesis using DNA nanoball-patterned arrays. Cell 185, 1777–1792.e1721 (2022).

34. Zhang, M.J., et al. Polygenic enrichment distinguishes disease associations of individual cells in single-cell RNA-seq data. Nature Genetics 54, 1572–1580 (2022).

35. Joseph, S.B., Castrillo, A., Laffitte, B.A., Mangelsdorf, D.J. & Tontonoz, P. Reciprocal regulation of inflammation and lipid metabolism by liver X receptors. Nature Medicine 9, 213–219 (2003).

36. Levine, M.E., et al. An epigenetic biomarker of aging for lifespan and healthspan. Aging (Albany NY) 10, 573–591 (2018).

37. Fong, S., et al. Principal component-based clinical aging clocks identify signatures of healthy aging and targets for clinical intervention. Nature Aging 4, 1137–1152 (2024).

38. Lu, A.T., et al. DNA methylation GrimAge strongly predicts lifespan and healthspan. Aging (Albany NY) 11, 303–327 (2019).

39. Balachandran, A., et al. Pace of Aging analysis of healthspan and lifespan in older adults in the US and UK. Nature Aging 5, 1132–1142 (2025).

40. Sehgal, R., et al. Systems Age: a single blood methylation test to quantify aging heterogeneity across 11 physiological systems. Nature Aging 5, 1880–1896 (2025).

41. Zhang, S., et al. A metabolomic profile of biological aging in 250,341 individuals from the UK Biobank. Nature Communications 15, 8081 (2024).

42. Patel, S., et al. GDF15 Provides an Endocrine Signal of Nutritional Stress in Mice and Humans. Cell Metabolism 29, 707–718.e708 (2019).

43. Moon, J.S., et al. Growth differentiation factor 15 protects against the aging-mediated systemic inflammatory response in humans and mice. Aging Cell 19, e13195 (2020).

44. Ritchie, S.C., et al. The Biomarker GlycA Is Associated with Chronic Inflammation and Predicts Long-Term Risk of Severe Infection. Cell Systems 1, 293–301 (2015).

45. Youm, Y.H., et al. Canonical Nlrp3 inflammasome links systemic low-grade inflammation to functional decline in aging. Cell Metabolism 18, 519–532 (2013).

46. Basisty, N., et al. A proteomic atlas of senescence-associated secretomes for aging biomarker development. PLoS Biology 18, e3000599 (2020).

47. Pyrkov, T.V., et al. Longitudinal analysis of blood markers reveals progressive loss of resilience and predicts human lifespan limit. Nature Communications 12, 2765 (2021).

48. Mutz, J., Iniesta, R. & Lewis, C.M. Metabolomic age (MileAge) predicts health and life span: A comparison of multiple machine learning algorithms. Science Advances 10, eadp3743 (2024).

49. Duan, T., et al. NGBoost: Natural Gradient Boosting for Probabilistic Prediction. Proceedings of the 37th International Conference on Machine Learning 119, 2690–2700 (2020).

50. März, A. & Kneib, T. Distributional gradient boosting machines. arXiv preprint arXiv:2204.00778 (2022).

51. März, A. XGBoostLSS--An extension of XGBoost to probabilistic forecasting. arXiv preprint arXiv:1907.03178 (2019).

52. Kuo, C.L., et al. A proteomic signature of healthspan. PNAS 122, e2414086122 (2025).

53. Zenin, A., et al. Identification of 12 genetic loci associated with human healthspan. Communications Biology 2, 41 (2019).

54. Barron, E., et al. Association between the English National Health Service Diabetes Prevention Programme and incident multiple long-term conditions. Nature Medicine 31, 3825–3831 (2025).

55. Mbatchou, J., et al. Computationally efficient whole-genome regression for quantitative and binary traits. Nature Genetics 53, 1097–1103 (2021).

56. Willer, C.J., Li, Y. & Abecasis, G.R. METAL: fast and efficient meta-analysis of genomewide association scans. Bioinformatics 26, 2190–2191 (2010).

57. Watanabe, K., Taskesen, E., van Bochoven, A. & Posthuma, D. Functional mapping and annotation of genetic associations with FUMA. Nature Communications 8, 1826 (2017).

58. Auton, A., et al. A global reference for human genetic variation. Nature 526, 68–74 (2015).

59. Bulik-Sullivan, B.K., et al. LD Score regression distinguishes confounding from polygenicity in genome-wide association studies. Nature Genetics 47, 291–295 (2015).

60. Grotzinger, A.D., et al. Genomic structural equation modelling provides insights into the multivariate genetic architecture of complex traits. Nature Human Behaviour 3, 513–525 (2019).

61. Bulik-Sullivan, B., et al. An atlas of genetic correlations across human diseases and traits. Nature Genetics 47, 1236–1241 (2015).

62. GTEx Consortium. The GTEx Consortium atlas of genetic regulatory effects across human tissues. Science 369, 1318–1330 (2020).

63. Chang, C.C., et al. Second-generation PLINK: rising to the challenge of larger and richer datasets. Gigascience 4, 7 (2015).

64. Song, L., Chen, W., Hou, J., Guo, M. & Yang, J. Spatially resolved mapping of cells associated with human complex traits. Nature 641, 932–941 (2025).

65. Consortium, T.S. Tabula Sapiens 2.0: A comprehensive transcriptomic atlas of human cell types. Cell 189, 7212–7227.e7218 (2026).

66. Kundaje, A., et al. Integrative analysis of 111 reference human epigenomes. Nature 518, 317–330 (2015).

67. Zhu, Z., et al. Integration of summary data from GWAS and eQTL studies predicts complex trait gene targets. Nature Genetics 48, 481–487 (2016).

68. Wu, Y., et al. Integrative analysis of omics summary data reveals putative mechanisms underlying complex traits. Nature Communications 9, 918 (2018).

69. Kuleshov, M.V., et al. Enrichr: a comprehensive gene set enrichment analysis web server 2016 update. Nucleic Acids Research 44, W90–97 (2016).

70. Piñero, J., et al. DisGeNET: a comprehensive platform integrating information on human disease-associated genes and variants. Nucleic Acids Research 45, D833–d839 (2017).

